# Contrasting Patterns of Cortical Volume Change in First-Episode Schizophrenia-Spectrum Psychosis

**DOI:** 10.64898/2026.10.08.26365057

**Authors:** Filip Spaniel, Antonin Skoch, Marian Kolenic, David Tomecek, Ivana Kawikova, Eduard Bakstein, Jaroslav Hlinka, Marketa Fialova, Lea Jakob, Vaclav Capek

## Abstract

**Importance:** Average gray matter volume (GMV) loss in early psychosis may conceal clinically relevant differences in brain change.

**Objective:** To identify longitudinal GMV-change patterns, their cross-site correspondence, and clinical associations.

**Design, Setting, and Participants:** Prospective cohort with T1-weighted MRI acquired during 2011–2026 at 2 Czech sites; median follow-up, approximately 13 months. Participants were 222 patients with first-episode schizophrenia-spectrum psychosis and 178 healthy controls frequency-matched by site and baseline age.

**Exposure:** First-episode psychosis vs healthy control status.

**Main Outcomes and Measures:** Diagnosis-blind clustering by site of image-quality-corrected, subsequently annualized GMV change across 333 cortical parcels; longitudinal voxel-based morphometry (VBM); Global Assessment of Functioning and consensus 5-factor Positive and Negative Syndrome Scale negative-symptom scores.

**Results:** Among 400 participants (mean [SD] age, 28.2 [7.5] years), 208 (52.0%) were male. Each site independently yielded 3 clusters: C1 (near-stable GMV), C2 (pronounced decrease), and C3 (increase). Cross-site centroid correlations across 333 parcels were strong for C2 and C3 (Spearman ρ = 0.865 and 0.861); C1 correlation was weak (ρ = 0.138), as expected for near-zero profiles with little regional variation. C2/C3 correspondence supported pooling matched site-specific clusters for post hoc analyses. Pooled clusters contained 77 patients and 125 controls in C1, 66 patients and 16 controls in C2, and 79 patients and 37 controls in C3. Suffixes −P and −HC identify patient and control subgroups. To situate patient changes relative to controls, post hoc VBM revealed an unanticipated gradient of increasingly pronounced GMV reduction across C1-HC → C1-P → C2-P, prominently involving association cortex, and bilateral ventral temporal GMV increase in C3-P (familywise error–corrected P < .05). The opposite contrast along this ordering showed periventricular changes consistent with progressively greater ventricular enlargement. C2-P had the lowest mean follow-up functioning and highest mean negative-symptom severity (adjusted omnibus P = .005 and .02); clinical changes from baseline did not differ significantly among patient groups.

**Conclusions and Relevance:** Early GMV change in psychosis was not uniform but comprised two contrasting directions: a clinically relevant, control-anchored pattern of increasingly pronounced GMV loss involving default mode regions and a distinct, directionally opposing ventral temporal increase.

**Key Points:** *Question:* What patterns of cortical gray matter volume change emerge over approximately 1 year after first-episode schizophrenia-spectrum psychosis?

*Findings:* In this 2-site cohort of 222 patients and 178 controls, nearly two-thirds of patients lay along a gradient from near-stability in controls to widespread volume decrease, whereas one-third showed an opposing ventral temporal increase. Follow-up functioning and negative symptoms differed significantly among patient groups, with the least favorable scores in those showing the greatest decrease.

*Meaning:* Directions of within-person brain change may capture clinically relevant heterogeneity obscured by average volume loss.

## Introduction

Schizophrenia-spectrum disorders vary in symptoms, course, functioning, treatment response, genetic liability, and neurobiology.^1–5^ Candidate psychosis biotypes and neuroanatomical subgroups nevertheless derive largely from cross-sectional data.^6–9^ Such subgroups capture between-person anatomy at one time point without distinguishing stable differences from distinct within-person directions or rates of change.^10I^

Longitudinal MRI studies report average gray matter volume (GMV) reduction, especially early in illness, with variable magnitude, distribution, and persistence; pronounced progression is not universal.^11–16^ Repeated imaging near onset may identify divergence before prolonged illness and treatment effects accumulate.^17–18^ Continuum and normative models further suggest some variation may extend dimensionally into the healthy population rather than define mutually exclusive diagnostic categories.^19–20^

Clustering participants by longitudinal GMV change can distinguish varying trajectories and may reveal differences in the magnitude, regional distribution, or direction of structural change. We asked whether early psychosis is characterized by a common pattern of brain change or by distinct, reproducible trajectories. To determine whether patient trajectories extend normative variation or represent qualitatively different patterns, we jointly clustered patients and healthy controls within each MRI site based on approximately 1-year cortical volume change across 333 brain regions, without diagnostic labels. We analyzed the 2 site-specific samples independently, then established cross-site correspondence of the resulting patterns from their cluster centroids before pooling matched clusters. Post hoc longitudinal voxel-based morphometry then characterized these patterns at finer spatial resolution, and clinical analyses examined whether patient groups differed in symptoms and functioning.

## Methods

### Participants and Study Design

This prospective cohort came from the Early-Stage Schizophrenia Outcome study, which began recruitment in 2011. We recruited patients through 15 inpatient psychiatric facilities and controls through community advertisements. The first episode was operationalized as the first hospitalization for psychosis.^21–22^ Of 573 patients and 317 controls with baseline imaging, 329 and 220, respectively, completed follow-up. After eligibility and MRI quality-control screening, we analyzed 222 patients with first-episode schizophrenia-spectrum psychosis and a schizophrenia-spectrum diagnosis confirmed at V2, along with 178 controls (eFigure 1 in Supplement).

**Figure 1.**
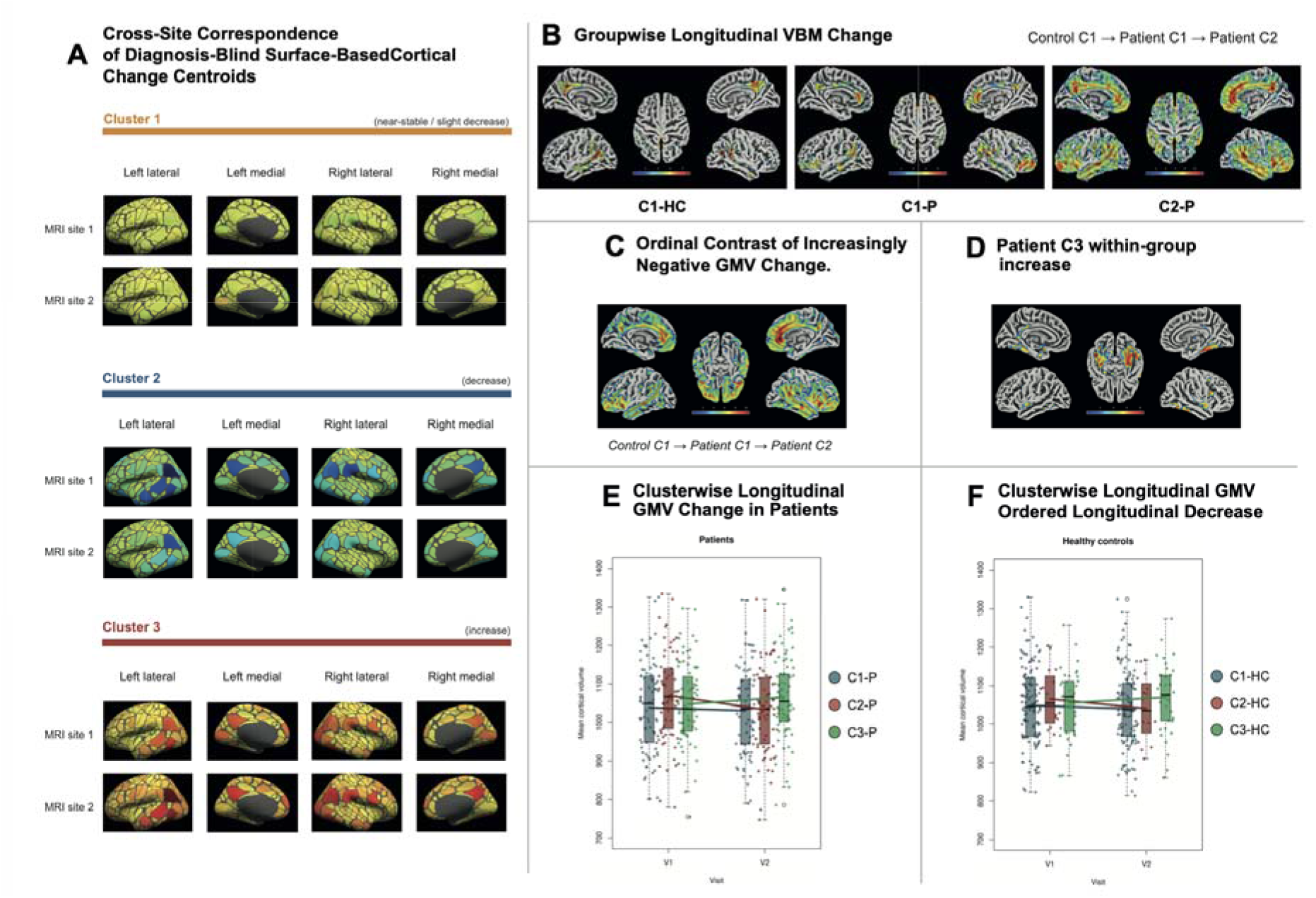
Cross-Site Correspondence and Voxelwise Characterization of Longitudinal Cortical Volume-Change Clusters. **A**, Mean cortical volume-change centroids after image-quality correction of raw V2−V1 change and subsequent annualization across 333 Gordon atlas parcels for the 3 diagnosis-blind clusters identified independently at MRI sites 1 and 2 and aligned as C1 (near-stable/slight decrease), C2 (pronounced decrease), and C3 (increase). Cross-site correspondence was strong for C2 (Spearman ρ = 0.86) and C3 (ρ = 0.86). C1 centroids showed near-zero, low-dispersion profiles, rendering their rank correlation (ρ = 0.14) minimally informative. **B-D**, Post hoc longitudinal VBM in separate patient and control subgroups corresponding to these clusters. All findings were FWE-corrected at P < .05. **B**, Annualized gray matter volume-change maps for C1-HC (n = 125), C1-P (n = 77), and C2-P (n = 66), suggesting progressively more negative change along the C1-HC → C1-P → C2-P gradient. **C**, Ordinal contrast formally testing this ordered decrease, with widespread involvement of association cortex, including prominent default mode network regions. **D**, Within-group gray matter volume increase in C3-P (n = 79), centered bilaterally in ventral temporal cortex. **E and F**, Participant-level mean cortical volume across the 333 parcels at baseline (V1) and follow-up (V2), stratified by cluster in patients (**E**) and controls (**F**), providing an unthresholded view of the longitudinal directions represented by C1-C3. Colors indicate C1 (blue), C2 (red), and C3 (green).

Eligible patients were aged 18 to 60 years, had untreated psychosis for <12 months and illness duration ≤36 months, and met International Statistical Classification of Diseases and Related Health Problems, Tenth Revision criteria for schizophrenia-spectrum psychosis. The M.I.N.I. International Neuropsychiatric Interview supported diagnoses, which at least 2 trained psychiatrists independently reviewed.^23^ Exclusion criteria for patients were intellectual disability, organic or substance-related mental disorders, severe neurologic disease, significant head trauma, cerebrovascular disease, epilepsy, migraine, hypertension, or endocrine disease. Controls were additionally excluded for any personal psychiatric disorder or a first-degree family history of major psychiatric illness.

Principal baseline diagnoses were schizophrenia (46.8%), acute polymorphic psychotic disorder with (34.7%) or without (14.9%) symptoms of schizophrenia, and schizoaffective disorder (2.3%); follow-up diagnoses were schizophrenia (63.5%), F23-spectrum acute and transient psychotic disorders (28.4%), and schizoaffective disorder (8.1%). Most baseline imaging occurred during patients’ first hospitalization. Mean (SD) illness duration at imaging was 5.1 (4.8) months (median, 3.0; range, 1.0–21.0 months). We summarized negative symptoms using the consensus 5-factor Positive and Negative Syndrome Scale model, and assessed functioning with the Global Assessment of Functioning scale.^24–27^ Further recruitment, diagnostic, assessment, and treatment details appear in the eMethods and eTable 6 in the Supplement. The Prague Psychiatric Center (PPC) Research Ethics Committee approved the protocol in 2011 (No. 69/11); responsible ethics committees approved subsequent amendments. After PPC became the National Institute of Mental Health in 2015, its Research Ethics Board continued oversight. All participants provided written informed consent.

### MRI Acquisition, Cortical Change, and Quality Control

Participants underwent 3-T imaging on a Siemens Trio at site 1 or Siemens Magnetom Prisma at site 2; each participant’s visits used the same scanner and protocol. Mean (SD) interscan intervals were 13.8 (2.8) months in patients and 17.1 (9.5) in controls (P < .001) but did not differ across patient or control subgroups within C1–C3. We processed T1-weighted images with FreeSurfer 6.0 and divided visually inspected cortical surfaces into 333 Gordon atlas parcels.^28^ For each parcel, raw within-participant cortical volume change was first calculated as visit 2 cortical volume minus visit 1 cortical volume (V2−V1). We omitted Intracranial-volume (ICV) scaling for this within-participant change outcome.

Within-participant change from same-scanner, same-protocol longitudinal examinations reduces static cross-scanner offsets. We evaluated image quality using (i) ratings by the MRI operator performing the scan and (ii) the CAT12 integrated image quality rating ^29–34^. Because quantitative structural image-quality measures can covary with morphometric estimates,^31,35^ estimating the image-quality effect in patients could remove disease-associated and quality-related variance. Separately within each MRI site and for each cortical parcel, raw within-participant volume change (V2−V1) was modeled as a function of change in CAT12 image quality (dIQR) using healthy controls only. The fitted values from these control-derived models, which included an intercept, were subtracted from the raw V2−V1 change for both patients and healthy controls, yielding image-quality-corrected longitudinal volume-change residuals relative to the control-derived model. Only after this correction were the residualized changes divided by the V1–V2 interscan interval in years to obtain the annualized features used for site-specific clustering. Acquisition, processing, and quality-control details appear in eMethods in the Supplement.

### Exploratory Site-Specific Clustering

In each of the 2 nonoverlapping MRI-site samples, we independently applied hierarchical agglomerative clustering using Euclidean distance and Ward.D2 linkage to each participant’s 333-parcel annualized profile of image-quality-corrected change^36–39^. We entered patients and controls together without diagnosis labels. Both sites used the same analytical procedure, without either site’s information defining the other’s clustering solution. We used the Cattell-Nelson-Gorsuch scree criterion to determine the number of clusters. ^40^ Each site independently yielded three clusters (eFigure 2 in Supplement). Clustering implementation and complementary Gap-statistic analyses are detailed in the eMethods and eResults in the Supplement. We used the exploratory 3-cluster solution to match recurrent directions of change, not as evidence of three discrete biological subtypes.

### Cross-Site Correspondence and Pooling

Each site-specific cluster’s centroid was its mean 333-parcel annualized profile of image-quality-corrected change. Centroid-profile similarity aligned corresponding near-stable/slight-decrease (C1), pronounced-decrease (C2), and increase (C3) clusters across sites. We quantified cross-site correspondence using parcelwise Spearman correlations between corresponding centroids and, for C2 and C3, between direction vectors after subtracting the site’s C1 centroid. Direct centroid correlations used 5000-resample bootstrap 95% CIs. Pooling occurred only after evaluating cross-site correspondence exclusively from independently derived imaging centroids, without diagnosis or clinical outcomes; strong cross-site similarity of pronounced-decrease and increase patterns supported pooling corresponding site-specific assignments. Pooling neither contributed to cluster discovery nor redefined membership. This tested reproducibility of group-level change patterns rather than participant-level assignment stability. Diagnosis-specific subgroups C1-P to C3-P denote patients and C1-HC to C3-HC denote healthy controls within each pooled cluster.

### Pooled Voxelwise Analysis

Using matched site-specific assignments, post hoc longitudinal VBM in SPM12 tested whether 333-parcel surface-based cortical-change patterns appeared as coherent, FWE–corrected voxelwise GMV change at another spatial resolution. Annualized participant-level GMV-change maps were normalized to Montreal Neurological Institute space, smoothed with an 8-mm full-width at half-maximum kernel, and analyzed with age and sex covariates. Visual inspection of separate groupwise maps suggested a gradient of increasingly negative GMV change from C1□HC through C1□P to C2□P. In subsequent post hoc group-level analysis, an ordinal regressor coded C1□HC, C1□P, and C2□P as 0, 1, and 2, respectively; its negative contrast formally tested the apparent ordering of increasingly negative change. C3 P was analyzed separately for within-group GMV increase. C2-HC and C3-HC were retained descriptively but not modeled in the principal voxelwise contrasts, which focused on the negative-change pattern anchored to the larger C1-HC subgroup and the opposing increase pattern in C3-P. Significance thresholds were voxel-level FWE-corrected P < .05 and ≥50 contiguous voxels. This algorithmically distinct longitudinal morphometric pipeline provided anatomical characterization after cross-site matching, without aiming to revalidate exploratory clustering or participant-level assignments. Because both pipelines used the same T1-weighted examinations, their results were not statistically independent; we interpreted agreement in direction and topography as cross-pipeline, cross-resolution anatomical corroboration. Full processing and model details appear in eMethods in the Supplement. Separately, ventricular volume normalized to ICV at V1, V2, and V2−V1 change were compared across C1-P, C2-P, and C3-P using Kruskal-Wallis tests with Holm correction.

### Pooled Clinical Analysis

Among pooled patient subgroups (C1□P to C3□P), we compared global functioning and negative-symptom scores at baseline, follow-up, and follow-up-minus-baseline change using Kruskal-Wallis tests with epsilon squared effect sizes. Holm adjustment was separate for baseline, follow-up, and change scores. We tested twelve baseline demographic, illness, treatment, and global-anatomy variables for detectable imbalance, not equivalence (eTable 1 in Supplement). We repeated clinical comparisons by MRI site (eTable 2 in Supplement). Chi-square tests compared patient-control distributions across C1-C3. Tests were 2-sided (P < .05); available-case sample sizes are reported.

## Results

### Participants and Quality Control

The final sample comprised 222 patients and 178 controls (400 participants; 800 MRI examinations). Patients’ mean (SD) age was 28.1 (7.7) years; 127 (57.2%) were men. Patients and controls were similar in age but differed in sex distribution and educational attainment (eResults in the Supplement). Sites 1 and 2 included 142 and 258 participants, respectively, with nearly identical patient-control proportions; each participant’s longitudinal examinations used the same site and scanner. After control-derived correction, longitudinal change in CAT12 image quality showed no detectable residual association with mean cortical GMV change in patients (Spearman ρ = 0.03; P = .63; eTable 13 in the Supplement).

### Independent Clustering and Cross-Site Correspondence

Exploratory diagnosis-blind analysis combining patients and controls yielded 3 clusters independently per site. Cross-site parcelwise correlations were strong for C2 (pronounced-decrease) and C3 (increase) centroids (Spearman ρ = 0.865 and 0.861, respectively) but weak for C1 (near-stable) centroids (ρ = 0.138). Corresponding 5000-resample bootstrap estimates (95% CIs) were 0.801 (0.722 to 0.852), 0.781 (0.687 to 0.845), and 0.088 (-0.149 to 0.306), respectively. Relative to each site’s near-stable centroid, cross-site direction-vector correlations remained strong (C2 - C1, ρ = 0.823; C3 - C1, ρ = 0.812) (eTable 3 in Supplement).

Near-stable (C1) centroids were near zero with substantially less between-parcel dispersion, leaving little systematic spatial variation for rank-based comparison. Small compositional or measurement differences can therefore alter parcel ranks despite similarly low-amplitude overall change. Thus, low C1 correlation was less informative about near-stable-state cross-site correspondence, whereas C2 and C3 centroids had sufficient spatial variation for strong topographic correspondence.

After evaluating cross-site correspondence, we combined participants with matching site-specific C1-C3 labels for downstream analyses, retaining independently derived within-site assignments without pooled reclustering (eTable 4 in Supplement).

This yielded C1 (near-stable or slight decrease; n = 202), C2 (pronounced decrease; n = 82), and C3 (increase; n = 116). Their mean (SD) cortical volume changes after image-quality correction and subsequent annualization were −1.80 (6.90), −24.69 (15.06), and 19.30 (13.36) mm³/y, respectively (Table 1; eTable 16 in the Supplement). Patient-control distributions differed across C1-C3 (*P* < .001): C1 comprised 77 patients (C1□P; 34.7% of patients) and 125 controls (C1□HC; 70.2% of controls); C2 comprised 66 patients (C2□P; 29.7%) and 16 controls (C2□HC; 9.0%); and C3 comprised 79 patients (C3□P; 35.6%) and 37 controls (C3□HC; 20.8%). Although effect sizes differed by site, the C2□P subgroup had the least favorable mean follow-up functioning and negative-symptom scores at both sites after establishing imaging-based correspondence. Site-specific means (C1-P/C2-P/C3-P) were 83.5/68.2/82.7 and 79.4/73.0/75.2 for follow-up functioning and 1.9/2.6/2.0 and 1.7/2.1/2.1 for negative symptoms at sites 1 and 2, respectively. Baseline clinical differences were site-specific: absent at site 1 but present at site 2, where C3-P showed lower functioning and greater negative-symptom severity (eTable 2 in Supplement). eTable 5 in Supplement reports the secondary, exploratory comparison of combined C1-P and C2-P with C3-P. None of the 12 prespecified baseline patient variables differed across pooled patient subgroups C1-P to C3-P after Holm correction.

**Table 1.** Combined Site-Specific Image-Quality-Corrected Longitudinal Cortical Volume-Change Clusters and Participant Distribution.

| Group | Cortical volume change | All, No. | Corrected annualized change, mm<sup>3</sup>/y, mean (SD) | Corrected annualized change, mm<sup>3</sup>/y, median [IQR] | Patients, No. (%) | Controls, No. (%) |
| --- | --- | --- | --- | --- | --- | --- |
| C1 | Near-stable or slight decrease | 202 | -1.80 (6.90) | -1.60 [-6.03 to 3.26] | 77 (34.7) | 125 (70.2) |
| C2 | Pronounced decrease | 82 | -24.69 (15.06) | -22.25 [-31.78 to -15.17] | 66 (29.7) | 16 (9.0) |
| C3 | Increase | 116 | 19.30 (13.36) | 17.08 [11.49 to 21.64] | 79 (35.6) | 37 (20.8) |

Abbreviations: IQR, interquartile range; SD, standard deviation. C1, C2, and C3 are participant-level clusters containing both patients and controls. Percentages are calculated within diagnostic groups. Overall patient-control distribution, $P < .001$ . Corrected annualized change = [raw V2–V1 change – fitted value from the site- and parcel-specific healthy-control dIQR model] / interscan interval in years.

### Pooled Voxelwise Characterization

Separate groupwise maps visually suggested increasingly negative GMV change across C1-HC, C1-P, and C2-P. Formal testing of this apparent ordering showed widespread, increasingly negative GMV change across frontal, temporal, parietal, cingulate, insular, sensorimotor, occipital, subcortical, and cerebellar regions. Medial prefrontal, angular, precuneus, temporal, and cingulate areas overlapping the default mode network were prominent, although the map extended beyond that network (eTable 7 and eFigure 3 in Supplement).

At voxel-level FWE–corrected *P* < .05 with ≥50 voxels, the C1□HC–C1□P–C2□P ordered-decrease contrast produced a widespread cluster of 118□739 voxels (peak *T* = 10.65; Montreal Neurological Institute coordinates, 4, 38, −2). The reverse C1□HC–C1□P–C2□P contrast identified only 1 cluster meeting the extent criterion (106 voxels; peak *T* = 6.38) confined to midline periventricular regions, anatomically compatible with progressively greater ventricular enlargement across C1□HC–C1□P–C2□P (eTable 8 and eFigure 4 in the Supplement). Separately, ventricular-volume normalized to ICV change differed across C1-P, C2-P, and C3-P (0.0002, 0.0010, −0.0006; ε²=.339; Holm-adjusted P<.001; eTable 15 and eFigure 9 in the Supplement). eTables 9-11 and eFigures 5-7 in the Supplement show separate within-group decrease maps for C1-HC, C1-P, and C2-P.

Conversely, C3□P showed significant within-group increase centered in bilateral ventral temporal cortex, including fusiform, inferior temporal, and parahippocampal regions, with additional cortical, subcortical, and cerebellar involvement (FWE-corrected P < .05) (Figure 1D; eTable 12 and eFigure 8 in the Supplement). Thus, after matching site-specific groups solely by independently derived surface-based centroid profiles, FWE-corrected longitudinal VBM revealed coherent anatomical organization at another spatial resolution: progressively greater expression of a widespread negative-change pattern across C1-HC, C1-P, and C2-P, and a directionally opposing, bilateral ventral temporal–centered increase pattern in C3-P.

### Pooled Clinical Findings

Exploratory pairwise comparisons showed higher baseline functioning in C1□P than C3□P and greater baseline negative-symptom severity in C3□P than C1□P, but corresponding omnibus baseline findings did not survive adjustment.

At follow-up, functioning differed across C1□P, C2□P, and C3□P (adjusted P = .005; epsilon squared = .0621), with means of 81.1, 72.0, and 78.4; C2□P scored lower than C1□P and C3□P. Negative symptoms also differed (adjusted P = .02; epsilon squared = .0488), with means of 1.79, 2.25, and 2.05; C2-P had greater severity than C1□P. After Holm adjustment, interscan changes in functioning and negative-symptom severity did not differ among patient subgroups (both adjusted *P* = .12) (Table 2). Follow-up status, rather than adjusted between-subgroup clinical-change differences, was therefore the principal clinical signal. Additional findings appear in eResults in Supplement.

**Table 2.**
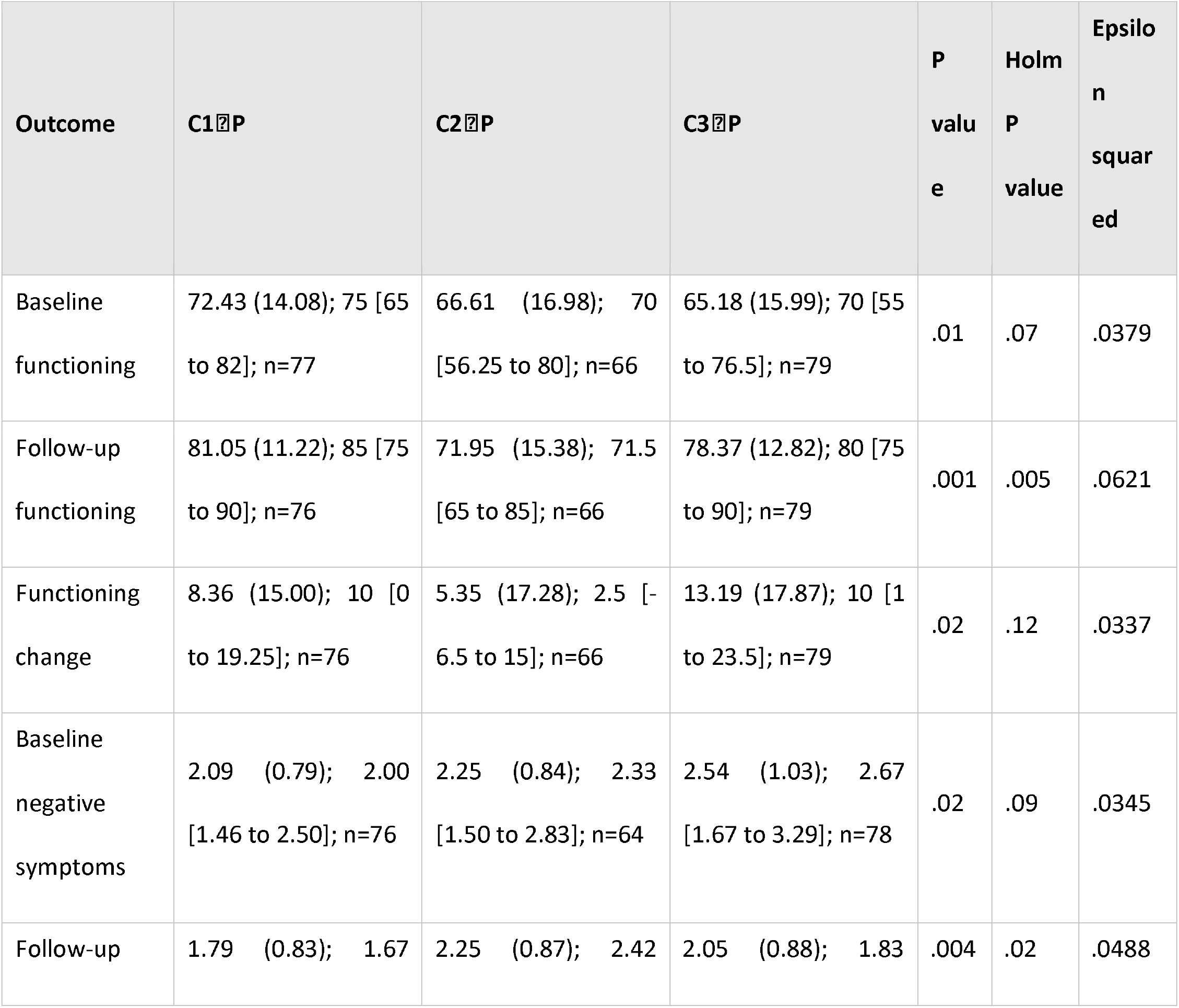
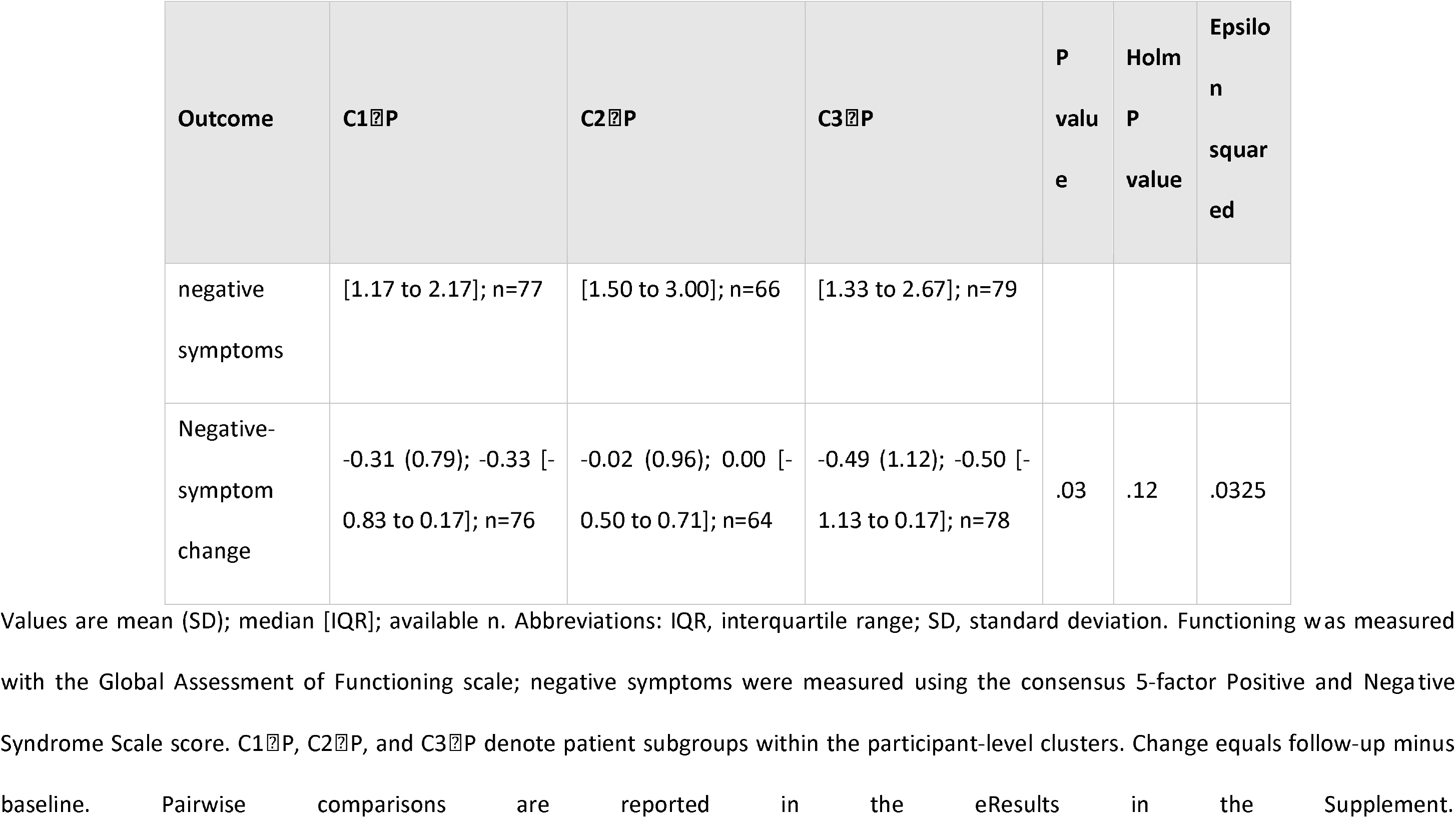
Global Functioning and Negative Symptoms Across Patient Subgroups Within C1-C3.

| Outcome | C1 $\bar{x}$ P | C2 $\bar{x}$ P | C3 $\bar{x}$ P | P value | Holm P value | Epsilon squared |
| --- | --- | --- | --- | --- | --- | --- |
| Baseline functioning | 72.43 (14.08); 75 [65 to 82]; n=77 | 66.61 (16.98); 70 [56.25 to 80]; n=66 | 65.18 (15.99); 70 [55 to 76.5]; n=79 | .01 | .07 | .0379 |
| Follow-up functioning | 81.05 (11.22); 85 [75 to 90]; n=76 | 71.95 (15.38); 71.5 [65 to 85]; n=66 | 78.37 (12.82); 80 [75 to 90]; n=79 | .001 | .005 | .0621 |
| Functioning change | 8.36 (15.00); 10 [-0 to 19.25]; n=76 | 5.35 (17.28); 2.5 [-6.5 to 15]; n=66 | 13.19 (17.87); 10 [-1 to 23.5]; n=79 | .02 | .12 | .0337 |
| Baseline negative symptoms | 2.09 (0.79); 2.00 [1.46 to 2.50]; n=76 | 2.25 (0.84); 2.33 [1.50 to 2.83]; n=64 | 2.54 (1.03); 2.67 [1.67 to 3.29]; n=78 | .02 | .09 | .0345 |
| Follow-up | 1.79 (0.83); 1.67 | 2.25 (0.87); 2.42 | 2.05 (0.88); 1.83 | .004 | .02 | .0488 |

| Outcome | C1 P | C2 P | C3 P | P<br>valu<br>e | Holm<br>P<br>value | Epsilo<br>n<br>squar<br>ed |
| --- | --- | --- | --- | --- | --- | --- |
| negative<br>symptoms | [1.17 to 2.17]; n=77 | [1.50 to 3.00]; n=66 | [1.33 to 2.67]; n=79 |  |  |  |
| Negative-symptom<br>change | -0.31 (0.79); -0.33 [-<br>0.83 to 0.17]; n=76 | -0.02 (0.96); 0.00 [-<br>0.50 to 0.71]; n=64 | -0.49 (1.12); -0.50 [-<br>1.13 to 0.17]; n=78 | .03 | .12 | .0325 |

Values are mean (SD); median [IQR]; available n. Abbreviations: IQR, interquartile range; SD, standard deviation. Functioning was measured with the Global Assessment of Functioning scale; negative symptoms were measured using the consensus 5-factor Positive and Negative Syndrome Scale score. C1 P, C2 P, and C3 P denote patient subgroups within the participant-level clusters. Change equals follow-up minus baseline. Pairwise comparisons are reported in the eResults in the Supplement.

## Discussion

In this 2-site cohort study, diagnosis-blind clustering of patients and controls independently identified low-change, pronounced-decrease, and increase profiles, with strong cross-site correspondence for the latter 2. Post hoc longitudinal VBM revealed increasingly negative GMV change across C1-HC, C1-P, and C2-P and bilateral ventral temporal–centered increase in C3-P. These findings support an organization of early structural change into an ordered negative-change pattern and an opposing increase pattern.

These findings extend longitudinal evidence of average GMV reduction and heterogeneous progression in psychosis.^11–16^ Coexisting directions of change may contribute to differences between study-level averages, consistent with broader accounts of cortical reorganization during early illness.^17,18,39^

Complementing cross-sectional biotypes and anatomical subgroups,^6–9^ grouping by within-person change distinguishes ongoing change from between-person anatomical differences.^10^ Controls occurred in all 3 combined groups; these profiles should therefore not be equated with disease-specific biological subtypes.

The C1-HC–C1-P–C2-P contrast characterized the negative-change component relative to the largest control subgroup; it did not order the full sample. Near-stability described residualized mean change, not absence of regional decreases. The reverse contrast yielded restricted periventricular signals. Separate patient-only volumetry showed greatest ventricular expansion in C2-P, little change in C1-P, and decrease in C3-P.

Shared anatomy may reflect amplification of processes involved in normative remodeling, which is regionally heterogeneous.^41–43^ Brain-age studies provide related context,^44–47^ including a cross-sectional ENIGMA study reporting a 3.55-year higher brain-predicted age difference in schizophrenia.^48^ However, we did not estimate brain age or normative age slopes; accelerated aging cannot be inferred. Prominent association regions overlapped the default mode network, consistent with literature on the vulnerability of highly connected, metabolically demanding, late-maturing cortex.^49–58^ The map extended beyond this network, and descriptive overlap does not establish network selectivity or propagation.

C3-P highlights a direction of change obscured by average GMV loss. Its ventral temporal increase does not establish compensation or recovery: T1-weighted morphometry is sensitive to tissue contrast, myelination, segmentation, and extracellular water.^59–66^ Medication, maturation, inflammatory or fluid-related changes, and measurement variation may also contribute. Within-group increase does not establish differences from controls or the decrease groups, which were not directly tested voxelwise.

C2-P had the least favorable mean follow-up functioning and negative-symptom scores, consistent with associations between brain measures and clinical status.^67–69^ Baseline omnibus differences and differences in clinical change did not survive multiplicity adjustment. Clinical relevance therefore concerned follow-up status. Because group assignment used both examinations, prognostic utility requires independent validation against subsequent outcomes.

### Limitations

Two examinations cannot resolve nonlinear trajectories, and annualization does not eliminate potential bias from unequal follow-up intervals. Attrition may introduce selection bias. Cluster number was sensitive to image-quality correction: an all-participant sensitivity analysis yielded 4 clusters at site 1 and 3 at site 2, although the principal uncorrected change directions persisted under fixed assignments (eTables 13-14 in the Supplement). C1 topographic correspondence was weak, site 1 C2 was small, and participant-level assignment stability and external replication remain untested. Pooled VBM did not adjust for site despite unequal site composition, leaving possible residual confounding; moreover, the clustering and VBM pipelines used the same scans and therefore do not constitute statistically independent validation. The selected post hoc ordinal contrast cannot establish a population continuum or equal spacing among groups. Finally, cumulative treatment exposure was unavailable, and T1-weighted morphometry cannot identify neuronal loss or distinguish among underlying biological mechanisms.

## Conclusions

Early brain structural change after first-episode psychosis followed no single trajectory. Instead, longitudinal cortical GMV variation followed two contrasting directions: an ordered, control-anchored gradient of increasingly pronounced widespread loss and a directionally opposing increase pattern. Less favorable clinical status in patients expressing the strongest GMV loss further indicates that this organization carries clinical information. These findings shift descriptions of early structural heterogeneity from fixed neuroanatomical subtypes toward distinct directions of within-person brain change.

## Data Availability

Data Sharing Statement
Data available: Yes.
Data types: Deidentified participant-level data and data dictionary.
Data shared: Participant ID, patient/control status, imaging site, cluster assignment, and image-quality-corrected annualized cortical-volume change across 333 parcels.
Supporting documents: Analytic code.
When available: With publication.
How to access data: Zenodo (DOI: https://doi.org/10.5281/zenodo.23211633).
Who can access the data: Anyone.
Types of analyses: Any research purpose.
Restrictions: Raw MRI and additional potentially identifying clinical data will not be publicly shared.

https://doi.org/10.5281/zenodo.23211633

## Article Information

### Author Contributions

Dr Spaniel had full access to all the data in the study and takes responsibility for the integrity of the data and the accuracy of the data analysis.

### Concept and design

Spaniel.

### Acquisition, analysis, or interpretation of data

Spaniel, Skoch, Kolenic, Tomecek, Bakstein, Hlinka, Capek.

### Drafting of the manuscript

Spaniel, Capek, Kolenic, Skoch, Bakstein, Kawikova.

### Critical review of the manuscript for important intellectual content

All authors.

### Statistical analysis

Capek.

### Obtained funding

Spaniel.

### Administrative, technical, or material support

Fialova, Jakob. Supervision: Spaniel.

### Data Sharing Statement

See the Supplement.

## Acknowledgment of AI Use

The authors used ChatGPT (GPT-5.6 Sol; OpenAI) in August and September 2026 for language editing, concision, and refinement of manuscript presentation. It was not used for study design, data analysis, or statistical analysis. All scientific content, results, interpretations, and references were independently reviewed and verified by the authors, who retain full responsibility for the manuscript.

