## Supplementary material for "Contrasting Patterns of Cortical Volume Change in First-Episode Schizophrenia-Spectrum Psychosis": SUPLLEMENTARY

**Contents**

**eMethods. Supplementary Methods**	**3**

**eResults. Supplementary Results**	**14**

eTable 1. Baseline Characteristics Across Combined Center-Specific Patient Pattern Groups	20

eTable 2. Center-Stratified Clinical Analyses	22

eTable 3. Cross-Center Similarity of Center-Specific Cluster Centroids	24

eTable 4. Imaging Site Composition of Combined Center-Specific Pattern Groups After Controls-Only Image-Quality Correction	26

eTable 5. Exploratory Clinical Comparison of Combined Patient Clusters 1 and 2 With Patient Cluster 3	27

eTable 6. Longitudinal Treatment and Compliance Indicators Across Combined Center-Specific Patient Pattern Groups	28

eFigure 1. Participant Flow Through the Study	29

eFigure 2. Site-Specific Cluster Dendrograms	30

eTable 7. Neuromorphometrics Atlas Labeling of Significant Clusters for the C1-HC–C1-P–C2-P Group Gradient Gray Matter Volume Decrease Contrast	32

eFigure 3. Ordered C1-HC–C1-P–C2-P Gray Matter Volume Decrease	34

eTable 8. Voxelwise Findings for the Reverse-Ordered C1-HC–C1-P–C2-P Group Gradient Gray Matter Volume Increase Contrast	35

eFigure 4. Reverse-Ordered C1-HC–C1-P–C2-P Gray Matter Volume Change	36

eTable 9. Neuromorphometrics Atlas Labeling of Significant Clusters for Within-Group Decrease in Annualized Gray Matter Volume in Control Cluster 1	37

eFigure 5. Gray Matter Volume Decrease in Control Cluster 1	39

eTable 10. Neuromorphometrics Atlas Labeling of Significant Clusters for Within-Group Decrease in Annualized Gray Matter Volume in Patient Cluster 1	40

eFigure 6. Gray Matter Volume Decrease in Patient Cluster 1	43

eTable 11. Neuromorphometrics Atlas Labeling of Significant Clusters for Within-Group Decrease in Annualized Gray Matter Volume in Patient Cluster 2	44

eFigure 7. Gray Matter Volume Decrease in Patient Cluster 2	46

eTable 12. Neuromorphometrics Atlas Labeling of Significant Clusters for Within-Group Increase in Annualized Gray Matter Volume in Patient Cluster 3	47

eFigure 8. Gray Matter Volume Increase in Patient Cluster 3	49

eTable 13. Image-Quality Correction and Sensitivity Analyses	50

eTable 14. Uncorrected Cortical-Volume Change Sensitivity Checks	51

eTable 15. Ventricular Volume Normalized to Intracranial Volume Across Patient Pattern Groups	52

eFigure 9. Ventricular-Volume/ICV Change Across Patient Pattern Groups	53

eTable 16. Annualized Cortical-Volume Change by Imaging Center, Diagnosis, and Pattern Group	54

Data Sharing Statement	55

eReferences	56

*52*

**eMethods. Supplementary Methods**

**Participants, Recruitment, and Diagnostic Procedures**

The Early-Stage Schizophrenia Outcome (ESO) study is a prospective longitudinal study of first-episode schizophrenia-spectrum psychosis conducted at the National Institute of Mental Health, Czech Republic. Recruitment began in 2011. Researchers recruited patients through the ESO Patient Enrolment Network across 15 inpatient psychiatric facilities; controls were recruited through community advertisements. A first episode was operationalized as the first hospitalization for psychosis, consistent with previous imaging studies.^1-2^ Project records did not retain a complete screening denominator; approximately 1 in 3 screened patients was enrolled.

At dataset closure in April 2026, 573 patients and 317 healthy controls had completed V1 MRI. 329 patients and 220 controls completed V2 MRI; therefore, 244 patients and 97 controls with V1 MRI did not complete V2 MRI. Among the 329 patients who completed V2, 76 were excluded because the duration of untreated psychosis (DUP) was 12 months or longer, 3 because the final diagnosis was outside the prespecified schizophrenia-spectrum diagnoses, and none because illness duration exceeded 36 months. No additional patients were excluded for age, intellectual disability, severe neurologic disease, significant head trauma or intracranial hemorrhage, substance dependence, other eligibility criteria, or missing non-MRI variables required for the primary analysis. We further excluded 28 unique patients during MRI/image-quality control, yielding 222 patients in the final analytic sample. MRI-quality-control failures were overlapping and included unusable V1 images in 12 participants, unusable V2 images in 18, and failed visual cortical-surface quality control in 15; these category counts therefore do not sum to 28.

Among the 220 controls who completed V2, 2 were excluded for psychiatric history, 1 for substance abuse, 1 for a first-degree relative with major psychiatric illness, 2 for neurologic/intellectual/head-trauma criteria, and 5 for MRI/image-quality failure. Of the remaining 209 eligible controls, we frequency-matched 178 to patients by site and baseline age for the final analytic sample.

Diagnoses were assigned using International Statistical Classification of Diseases and Related Health Problems, Tenth Revision (ICD-10) criteria, independently reviewed by at least 2 experienced psychiatrists, and supported by the Mini International Neuropsychiatric Interview administered at V1.^3,4^ Among the 222 patients, diagnoses at V1 were schizophrenia (F20.x; n = 104 [46.8%]), acute polymorphic psychotic disorder with symptoms of schizophrenia (F23.1; n = 77 [34.7%]), acute polymorphic psychotic disorder without symptoms of schizophrenia (F23.0; n = 33 [14.9%]), acute schizophrenia-like psychotic disorder (F23.2; n = 2 [0.9%]), other acute predominantly delusional psychotic disorder (F23.3; n = 1 [0.5%]), and schizoaffective disorder (F25.x; n = 5 [2.3%]). At follow-up, 141 patients (63.5%) were diagnosed with schizophrenia, 45 (20.3%) with F23.1, 10 (4.5%) with F23.0, 5 (2.3%) with F23.2, 1 (0.5%) with F23.3, 2 (0.9%) with another F23.x acute and transient psychotic disorder, and 18 (8.1%) with schizoaffective disorder.

The DUP restriction reduced variation in untreated illness before baseline imaging; longer duration has been associated with lower regional gray matter volume or density.^5-7^ Exclusion criteria for patients were intellectual disability, organic or substance-related mental disorders, severe neurologic disease, significant head trauma, cerebrovascular disease, epilepsy, migraine, hypertension, or endocrine disease. Controls were additionally excluded for any personal psychiatric disorder or a first-degree family history of major psychiatric illness.

**Clinical Assessments and Treatment Ascertainment**

We assessed psychopathology with the Positive and Negative Syndrome Scale (PANSS).^8^ Negative symptoms were summarized using the Wallwork consensus 5-factor model,^9-10^ with the negative factor defined by PANSS items N1 (blunted affect), N2 (emotional withdrawal), N3 (poor rapport), N4 (passive/apathetic social withdrawal), N6 (lack of spontaneity and flow of conversation), and G7 (motor retardation). The reported PANSS-N value was the mean item score across these 6 items, retaining the 1-to-7 PANSS metric, with higher values indicating greater negative-symptom severity. We assessed functioning with the Global Assessment of Functioning (GAF) scale.^11^ Illness duration was established from records and collateral information from treating clinicians and family members. DUP was defined as the interval from psychosis onset to antipsychotic treatment initiation and was estimated from records, clinician reports, and patient interviews.^12^ Psychosis onset required at least 1 week of core positive symptoms; estimates were updated prospectively when new information became available.^13^

We recorded antipsychotic treatment at each imaging visit, including agent, prescribed dose, chlorpromazine-equivalent (CPZ-equivalent) dose, treatment duration, and medication adherence. Mean (SD) CPZ-equivalent dose was 379.1 (225.2) mg/d at V1 (n = 219) and 219.1 (191.1) mg/d at V2 (n = 204). Olanzapine, risperidone, and aripiprazole were most frequently recorded at V1; aripiprazole, olanzapine, and quetiapine were most frequent at V2. Continuous treatment histories were unavailable, so we could not reconstruct cumulative exposure. Antipsychotic doses were converted to CPZ-equivalent doses using the method of Andreasen et al.^14^ For newer antipsychotics not covered by that framework, including paliperidone, lurasidone, cariprazine, and brexpiprazole, equivalence was supplemented using the Second International Consensus Study of Antipsychotic Dosing (ICSAD-2).^15^ Long-acting injectable antipsychotics were converted using published LAI dose-equivalence data, with injection doses expressed relative to their dosing interval when daily exposure was required.^16^ For concurrent antipsychotic treatment, the CPZ-equivalent doses of individual agents were summed to obtain total daily CPZ-equivalent dose.

We assessed medication adherence at both imaging visits with a 1-item, 7-point Hayward/Kemp compliance rating, with higher scores indicating greater treatment acceptance/adherence.^17-18^ Ratings integrated the clinical record, clinician assessment, and patient interview. Treatment analyses used available-case data without imputation. CPZ-equivalent dose was available for 219 patients at V1 and 204 at V2, and compliance ratings were available for 221 patients at V1 and 215 at V2. Analyses of change in named antipsychotic agent were restricted to the 213 patients with agent information available at both visits. Missing treatment observations were not imputed.

**Magnetic Resonance Imaging Acquisition**

Participants underwent imaging at 2 centers: site 1 (MRI SITE 1) used a 3-T Siemens Trio scanner, and site 2 (MRI SITE 2) used a 3-T Siemens Magnetom Prisma scanner (Siemens Healthineers). Each participant completed both longitudinal examinations on the same scanner using the same site-specific acquisition protocol. The morphometric analyses reported in this study used high-resolution sagittal T1-weighted 3-dimensional magnetization-prepared rapid gradient-echo (MPRAGE) images acquired at both visits.

At site 1, the 3-T Siemens Trio was equipped with a 12-channel volume head coil. The T1-weighted sagittal MPRAGE sequence used an echo time (TE) of 4.63 ms, repetition time (TR) of 2300 ms, inversion time (TI) of 900 ms, iPAT acceleration factor of 2, 1 x 1 x 1 mm spatial resolution, and an acquisition time of 5 minutes 30 seconds. The site 1 structural protocol also included a transverse 2-dimensional T2-weighted BLADE sequence (TE, 128 ms; TR, 5850 ms; resolution, 0.6 x 0.6 x 3.0 mm; acquisition time, 2 minutes 22 seconds) and a sagittal 3-dimensional T2-weighted FLAIR sequence (TE, 318 ms; TR, 6200 ms; TI, 2200 ms; iPAT, 2; resolution, 1 x 1 x 1 mm; acquisition time, 6 minutes 38 seconds).

At site 2, the 3-T Siemens Magnetom Prisma was equipped with a 64-channel receive head coil. The T1-weighted sagittal MPRAGE sequence used a TE of 2.34 ms, TR of 2400 ms, TI of 1000 ms, iPAT acceleration factor of 2, 0.7 x 0.7 x 0.7 mm spatial resolution, and an acquisition time of 7 minutes 40 seconds. The site 2 structural protocol also included a transverse 2-dimensional T2-weighted BLADE sequence (TE, 106 ms; TR, 5500 ms; iPAT, 2; resolution, 0.7 x 0.7 x 3.0 mm; acquisition time, 1 minute 30 seconds) and a sagittal 3-dimensional T2-weighted SPACE sequence (TE, 564 ms; TR, 3200 ms; iPAT, 2; resolution, 0.7 x 0.7 x 0.7 mm; acquisition time, 7 minutes 55 seconds).

No relevant scanner hardware, software, or T1 acquisition-protocol change was reported during the study period.

Site 1 Siemens Trio ran software version VB17A, and site 2 Siemens MAGNETOM Prisma ran syngo MR software version VE11B.

The protocol prespecified an approximately 12-month interval between V1 and V2. For patients, the mean (SD), median, and range between visits were 13.83 (2.79), 13.06, and 10.32 to 27.83 months; for controls, 17.06 (9.48), 13.96, and 9.56 to 87.16 months (P < .001). The longer upper tail among controls arose because some controls had originally participated during the cross-sectional phase at the beginning of ESO and were subsequently re-recruited when longitudinal follow-up was introduced. Variation in the interscan interval was handled by annualization; for the clustering analysis, the image-quality-corrected parcel changes were annualized only after the control-derived correction described below. Intervals did not differ across patient (P = .057) or control (P = .331) clusters. Among patients, the interval from illness onset to V1 was 5.12 (4.83) months (median, 3; range, 1-21 months) and did not differ across clusters (P = .7946).

**Surface-Based Morphometry**

T1-weighted images were processed with FreeSurfer version 6.0 using its longitudinal processing stream. We created a within-participant unbiased base template from the V1 and V2 examinations, then processed each time point longitudinally using information from the subject-specific template. Processing included intensity normalization, skull stripping, Talairach transformation, gray-white matter segmentation, cortical-surface reconstruction, and longitudinally informed surface estimation. We visually inspected each reconstruction and manually edited it when needed. We parceled cortical surfaces into 333 functionally defined regions using the Gordon atlas.^19^ Parcel labels were mapped to each participant's reconstructed cortical surface, and regional cortical volumes were extracted for V1 and V2. For the clustering analysis, no intracranial-volume adjustment was applied to parcel-wise cortical volumes. This decision reflected the within-participant change design: ICV is effectively stable within adults and cannot explain the direction of V2-minus-V1 change; baseline global GMV relative to ICV was evaluated separately as a patient-group characteristic. For each parcel, raw within-participant cortical-volume change was calculated as the V2-minus-V1 parcel-volume difference. This raw difference was first subjected to the control-derived image-quality correction described below; only the corrected difference was then divided by the interscan interval in years, yielding values in mm³/y. The resulting 333 image-quality-corrected annualized changes formed each participant's clustering feature vector.

**Hierarchical Clustering and Cluster-Number Selection**

Hierarchical agglomerative clustering based on Euclidean distance iteratively merged participants with similar 333-parcel image-quality-corrected annualized change profiles.^20-23^ Within each center, patients and controls were clustered together without diagnosis labels. We did not adjust the center-specific clustering features for age, sex, or diagnostic group, except for the CAT12 image-quality correction described below. No parcel values were missing in either center-specific clustering matrix. We implemented clustering in R version 4.3.3 using the dist and hclust functions from the stats package. We used Ward.D2 linkage for hierarchical clustering. We did not standardize the 333 image-quality-corrected annualized parcel features before calculating Euclidean distance. The number of clusters was not prespecified. We evaluated the number of clusters using complementary statistical criteria and independent center-specific analyses. We calculated the Cattell-Nelson-Gorsuch scree criterion from the height values returned by hclust using the nCng function from the nFactors package.^24^ As an additional clustering-specific diagnostic, we evaluated the Gap statistic for K = 2, 3, and 4 on the same Ward.D2 clustering within each center.

We selected the number of clusters independently in the 2 nonoverlapping imaging-center cohorts. The MRI SITE 1 cohort contained 142 participants and the MRI SITE 2 cohort 258. The Cattell-Nelson-Gorsuch criterion selected 3 clusters in both centers, and the Gap statistic peaked at K = 3 in both centers, although the differences between candidate solutions were modest. Thus, K = 3 was not interpreted as an unequivocally determined population structure, but as the solution receiving the most consistent support across the complementary clustering diagnostics and the 2 independent acquisition cohorts. After the parcel- and center-specific control-derived image-quality correction described below and subsequent annualization, Ward.D2 hierarchical clustering was performed separately in MRI SITE 1 and MRI SITE 2. Center-specific clusters were related across centers using centroid-profile similarity, and participants retained their center-specific assignments before being combined by corresponding pattern label for subsequent analyses. No participant-level clustering was performed across centers. These independent center analyses provided an internal cross-center robustness assessment but were not bootstrap/resampling stability analyses; we did not test stable individual assignments across centers.

**Cross-Center Centroid Correspondence Analysis**

To identify corresponding average cortical-change profiles across the 3-cluster solutions obtained independently at MRI SITE 1 and MRI SITE 2, we compared the native center-specific cluster centroids. For each cluster, the centroid was the arithmetic mean of the 333-parcel image-quality-corrected annualized cortical-volume change profile across all participants assigned to that cluster, irrespective of diagnosis. The analysis therefore used 3 MRI SITE 1 and 3 MRI SITE 2 centroid vectors derived after control-derived image-quality correction and subsequent annualization. We did not use clinical measures or patient-control composition to define the comparisons.

For each of the 9 MRI SITE 1-by-MRI SITE 2 centroid pairs, Spearman rank correlation and unscaled Euclidean distance were calculated across the 333 parcels. Spearman correlation summarized concordance of parcel ordering, whereas Euclidean distance summarized absolute separation in the native centroid feature space. Centroid-profile similarity was used to identify the corresponding pairs (MRI SITE 1 C1 with MRI SITE 2 C1, C2 with C2, and C3 with C3); in the distance matrix, each centroid's smallest cross-center distance was its corresponding counterpart. To evaluate the broader directions relative to the near-stable cluster, each center's C1 centroid was subtracted from its C2 and C3 centroids, and the resulting C2 - C1 and C3 - C1 vectors were correlated across centers using Spearman correlation.

We performed 5000 bootstrap resamples for each direct centroid Spearman correlation and provided a bootstrap central estimate with a 95% interval. These summaries were intended to quantify uncertainty in cross-center similarity of the average profiles and were kept separate from the observed correlations. No P values or multiplicity-adjusted tests were reported. We did not report bootstrap intervals for the direction-vector correlations. For each of the 5000 bootstrap iterations, subjects were resampled with replacement separately within each of the 6 native center-specific clusters (3 MRI SITE 1 and 3 MRI SITE 2 clusters); the 333-parcel centroid was recomputed as the mean across resampled subjects within each cluster, and Spearman correlations were recalculated for the resulting centroid pairs. Native cluster labels were held fixed, and clustering was not repeated. For each cell of the 5000 resulting correlation matrices, we reported the median as the bootstrap central estimate and the 2.5th and 97.5th percentiles as the 95% interval.

All 6 centroid vectors contained numeric values for all 333 parcels. The centroid analysis is an aggregate profile analysis and does not test whether individual participants would receive concordant assignments across centers or whether cluster membership is stable under resampling

**Image Quality Control and Control-Derived Correction**

In-scanner motion can bias automated cortical morphometry.^25-28^ Immediately after acquisition, operators rated overall T1-weighted image quality on a 4-point scale, with 4 indicating the greatest artifact burden; scans failing the prespecified threshold of >3 were excluded. CAT12 provided an integrated image quality rating (IQR) incorporating noise relative to tissue contrast, intensity inhomogeneity, and spatial resolution, with higher values indicating poorer quality.^29-30^ The exclusion threshold applied only to the operator quality rating; no threshold was applied to the CAT12 image-quality rating, which was treated as a continuous measure in the correction analyses.

Longitudinal change in the CAT12 integrated image-quality rating was defined as follow-up minus baseline (IQR_V2 - IQR_V1) and was not annualized. Because quantitative structural image-quality measures can covary with morphometric estimates,^27,31^ estimating this association from patient data could potentially remove disease-associated variance together with quality-related variance. We therefore estimated the association between longitudinal image-quality change and raw V2-minus-V1 parcelwise cortical-volume change exclusively in healthy controls, separately within each imaging center. We then subtracted the resulting parcel- and center-specific control-derived fitted values from the raw V2-minus-V1 changes of both patients and controls before annualization and center-specific clustering.

**Control-derived correction**

For each of the 333 Gordon atlas parcels, separately within each center, a controls-only linear regression modeled the unannualized V2-minus-V1 parcel-volume difference as the dependent variable, with longitudinal IQR change and an intercept as predictors. For both patients and controls, the corrected parcel-volume difference was calculated as the observed raw V2-minus-V1 difference minus the full control-derived model-predicted value, including the intercept, and only then annualized by dividing by the V1-to-V2 interval in years. Thus, the corrected values represent center- and parcel-specific residualized longitudinal change relative to the controls-derived model rather than raw anatomical change referenced to a literal zero. This approach allowed the image-quality association to vary across cortical regions and scanners while avoiding estimation of the nuisance relation from patient data used to characterize longitudinal heterogeneity. The subsequent center-specific clustering procedure was unchanged.

To assess whether the low-change, pronounced-decrease, and increase directions identified in the principal analysis could have been induced by the control-derived correction, we performed a post hoc descriptive analysis using annualized cortical-volume changes without CAT12 image-quality correction. The principal controls-only-corrected center-specific assignments were held fixed, and uncorrected mean annualized cortical-volume change was summarized after participants were combined by corresponding pattern label. This analysis assessed the direction of the underlying uncorrected morphometric signal and did not redefine the principal center-specific assignments or subsequent clinical analyses.

**All-participant IQR sensitivity correction**

A complementary, deliberately more aggressive sensitivity analysis estimated the image-quality-related component from the full patient-control sample. We fitted separate parcel-specific linear models for each of the 333 parcels, with parcel volume change as the dependent variable and longitudinal CAT12 image-quality rating change as the predictor. We repeated center-specific clustering using the resulting residualized longitudinal-change profiles. This analysis tested whether the principal findings persisted when a broader fraction of image-quality-associated variance was removed, including variance estimated from patients themselves

**Clinical and Statistical Analyses**

We examined center-stratified clusters for concordant patterns in global functioning and negative symptoms. After participants were combined across centers by corresponding center-specific pattern label, we compared these outcomes in patients at baseline, follow-up, and for follow-up-minus-baseline change. Positive functioning change and negative negative-symptom change indicated improvement. We used Kruskal-Wallis tests with epsilon-squared effect sizes and exploratory pairwise Wilcoxon rank-sum tests implemented with pairwise.wilcox.test from the stats package; we applied Holm adjustment separately to baseline, follow-up, and change-score families.

We performed the nonimaging analyses in R version 4.3.3 using the stats package.

We examined baseline comparability across patient clusters for age, sex, body mass index, cigarettes per day, education, diagnosis, age at psychosis onset, illness duration at V1, DUP, baseline CPZ-equivalent dose, medication compliance, and global gray matter volume normalized to intracranial volume. Continuous and ordinal variables used Kruskal-Wallis tests, sex used a chi-square test, and diagnosis used a Fisher exact test. We applied Holm adjustment across the full 12-comparison baseline family. We compared patient-control pattern distributions using chi-square tests. All tests were 2-sided with P < .05; variable-specific available-case sample sizes are reported, and no clinical or treatment observations were imputed.

We retained baseline compliance as the 12th prespecified baseline variable. Baseline compliance was available for 221 of 222 patients and did not differ across patient groups formed by combining corresponding center-specific pattern labels (P = .51). We therefore recalculated the Holm-adjusted P values in eTable 1 across all 12 baseline comparisons.

Medication variables did not define the clusters and were not covariates in primary voxelwise models. Antipsychotic exposure and illness duration, systemic inflammation, body mass index, and smoking have been associated with brain morphology.^32-37^ The measured treatment comparisons reduce concern about gross imbalance but do not exclude residual confounding from cumulative or time-varying exposure.

**Post Hoc Longitudinal Voxel-Based Morphometry**

We analyzed the same baseline and follow-up images in SPM12 (build 7771). Symmetric longitudinal registration included rigid-body alignment, intensity-inhomogeneity correction, and nonlinear within-participant deformation. This analysis constituted the principal high-resolution anatomical characterization of the cross-site–matched change patterns; it was not designed as an independent validation of the exploratory cluster assignments.

We segmented a participant-specific average image into gray matter, white matter, and cerebrospinal fluid; we then registered the gray- and white-matter segments across participants with DARTEL. Transformations normalized annualized gray matter change maps to Montreal Neurological Institute space.

Gray matter tissue maps were modulated by Jacobian determinants using the preserve amount option, smoothed with an 8-mm full-width at half-maximum Gaussian kernel, and restricted to the explicit mask brainmask_GMtight.nii. The explicit mask was the standard CAT12-distributed GM-tight mask in MNI152NLin2009cAsym space. We did not derive it from the study sample or any statistical contrast, and we applied no study-specific threshold; it was therefore independent of the tested effects. General linear models adjusted for age and sex. Control cluster 1 (n = 125), patient cluster 1 (n = 77), and patient cluster 2 (n = 66) entered an ordinal model (N = 268; 264 residual degrees of freedom) coded 0, 1, and 2. The negative contrast tested progressively greater decrease; the reverse contrast was inspected. We tested patient cluster 3 (n = 79) for within-group increase. Voxel-level family wise error (FWE) P < .05 and a 50-voxel extent threshold defined the reported findings.

Thresholded statistical parametric maps were anatomically labeled with the CAT12 Neuromorphometrics atlas. The eTables report cluster size, peak T value, peak Montreal Neurological Institute coordinates, and atlas regions comprising at least 1% of each cluster in the source export. Local resel-per-voxel values were used to account for nonstationarity of smoothness.

The ordered post hoc VBM analysis was restricted to control cluster 1, patient cluster 1, and patient cluster 2. We did not include patient cluster 3 in this trend analysis and examined it separately using a within-group longitudinal increase contrast. We did not perform a direct whole-brain patient C3-P vs C1-P/C2-P comparison.

**Direct Ventricular-Volume Analysis**

Ventricular volume was normalized to intracranial volume (ICV) at V1 and V2; V2-minus-V1 change was compared across C1-P, C2-P, and C3-P using Kruskal-Wallis tests with epsilon-squared effect sizes and Holm adjustment across V1, V2, and change.

**eResults. Supplementary Results**

**Patient-Control Baseline Characteristics**

At V1, patients had a mean (SD) age of 28.11 (7.67) years and controls 28.29 (7.38) years. Men comprised 127 of 222 patients (57.2%) and 81 of 178 controls (45.5%). Patient educational attainment was primary school, n = 40; vocational school without secondary graduation, n = 27; vocational school with secondary graduation, n = 14; secondary school, n = 85; and university, n = 56. Corresponding control counts were 5, 1, 6, 86, and 80.

**Diagnostic Follow-up and Transitions**

Individual diagnostic transitions from V1 to follow-up showed substantial stability among patients initially diagnosed with schizophrenia: 101 of 104 (97.1%) retained schizophrenia, while 3 (2.9%) received a follow-up diagnosis of schizoaffective disorder. Among 77 patients with F23.1 at V1, 41 (53.2%) retained F23.1, 26 (33.8%) were diagnosed with schizophrenia, 6 (7.8%) with schizoaffective disorder, 2 (2.6%) with F23.2, and 2 (2.6%) with another F23.x acute and transient psychotic disorder. Among 33 patients with F23.0 at V1, 10 (30.3%) retained F23.0, 13 (39.4%) were diagnosed with schizophrenia, 4 (12.1%) with F23.1, 4 (12.1%) with schizoaffective disorder, and 2 (6.1%) with F23.2. Of 2 patients with F23.2 at V1, 1 retained F23.2, and 1 was diagnosed with schizophrenia; the single patient with F23.3 and all 5 patients with schizoaffective disorder at V1 retained their respective categories. Overall, 159 of 222 patients (71.6%) retained the same reported ICD-10 diagnostic category and 63 (28.4%) changed category. F20 and F25 subcodes were collapsed within their respective diagnostic families.

**Center-Stratified Analyses**

The scree criterion identified 3 clusters independently in the MRI SITE 1 and MRI SITE 2 cohorts. At MRI SITE 1 (n = 142), center-specific clusters 1, 2, and 3 contained 81, 14, and 47 participants, respectively (patients/controls: 32/49, 14/0, and 33/14). At MRI SITE 2 (n = 258), the corresponding cluster sizes were 121, 68, and 69 (patients/controls: 45/76, 52/16, and 46/23). Baseline functioning and negative symptoms did not differ at MRI SITE 1; both differed at MRI SITE 2, where cluster 3 showed the least favorable baseline means. At follow-up, cluster 2 had the least favorable mean functioning and negative-symptom scores at both centers. Change patterns were directionally similar, but statistical strength varied (eTable 2). This parallel clinical ordering was observed after the imaging-based centroid correspondence was established. We evaluated cross-center correspondence of the center-specific average profiles using centroid analysis; we did not test stable individual assignments. As an additional clustering-specific validation, the Gap statistic was maximal at K = 3 in both centers (MRI SITE 1: K = 2, 0.5834; K = 3, 0.5861; K = 4, 0.5858; MRI SITE 2: K = 2, 0.9040; K = 3, 0.9074; K = 4, 0.8987).

**Cross-Center Centroid Correspondence Results**

The observed cross-center parcelwise correlations were strong for the pronounced-decrease C2 and increase C3 centroid pairs (Spearman ρ = .8646 and .8606, respectively) and weak for the near-stable C1 pair (ρ = .1377) (eTable 3). The complete 3 x 3 matrices showed that correspondingly numbered clusters were the closest cross-center pairs by Euclidean distance: 102.9909 for C1, 480.8695 for C2, and 251.3098 for C3.

The 5000-resample bootstrap yielded central estimates (95% intervals) of .0881 (-.1486 to .3056) for C1, .8005 (.7221 to .8520) for C2, and .7810 (.6867 to .8451) for C3. These values are reported separately from the observed correlations because the bootstrap central estimates did not equal the observed statistics; the bootstrap central estimate was the median of the 5000 resampled correlations, and the 95% interval was defined by the 2.5th and 97.5th percentiles. The slight exceedance of the upper bootstrap percentile by the observed C2 and C3 correlations can occur under this percentile-bootstrap construction.

The near-stable centroids were close to zero and had substantially lower between-parcel dispersion than the C2 and C3 centroids. Thus, the weak C1 rank correlation indicates that a reproducible parcel ordering was not demonstrated for this low-amplitude subcomponent. It does not by itself negate the strong C2 and C3 correspondence within the broader organization. Differences in C1 composition or variance, measurement variability, or other center-specific characteristics are plausible explanations, but the aggregate analysis does not distinguish among them.

When each center's C1 centroid was used as a low-change reference, the corresponding direction vectors remained strongly correlated across centers (C2 - C1, ρ = .8228; C3 - C1, ρ = .8122). Taken together, the centroid analysis supports cross-center correspondence of the pronounced-decrease and increase directions emerging from a central low-change state, rather than 3 equally coherent topographic phenotypes. It evaluates average profiles only and does not establish participant-level transfer, cluster-membership stability, or external replication.

**Quality-Correction Check**

CAT12 Image-Quality Distributions

At V1, mean (SD) CAT12 image-quality rating was 1.8793 (0.2131) in controls and 1.9331 (0.2491) in patients (raw P = .0415; Holm-adjusted P = .1176). At V2, corresponding values were 1.8830 (0.2306) and 1.9529 (0.2838) (raw P = .0392; Holm-adjusted P = .1176). Longitudinal change did not differ between groups: mean (SD) change was 0.0036 (0.2039) in controls and 0.0197 (0.2948) in patients (raw P = .9417; Holm-adjusted P = .9417). We tested patient-control differences at V1, V2, and in V2-minus-V1 change with 2-sided Mann-Whitney U tests, with Holm adjustment across these 3 comparisons. Controls had n = 178 for each comparison; 222 patients had nonmissing CAT12 observations.

**Results after controls-only correction**

After controls-only calibration, the scree procedure selected 3 clusters independently at MRI SITE 1 and MRI SITE 2. After center-specific clusters were related by centroid-pattern similarity, participants with corresponding labels were combined across centers for summary and downstream analyses. In these combined groups (N=400), C1 comprised 202 participants with mean (SD) image-quality-corrected annualized cortical-volume change of −1.80 (6.90) mm³/y; C2 comprised 82 participants with −24.69 (15.06) mm³/y; and C3 comprised 116 participants with 19.30 (13.36) mm³/y. Patient-control allocation remained strongly nonrandom (P<.001): C1 contained 125 controls and 77 patients, C2 contained 16 controls and 66 patients, and C3 contained 37 controls and 79 patients.

Image-quality correction and sensitivity results are summarized in eTable 13.

**Uncorrected-Data Sensitivity Check**

The directional organization observed in the principal analysis was already present in the cortical-volume data before image-quality correction.

To examine this directly, the controls-only-corrected center-specific assignments were held fixed and each participant’s corresponding cortical-volume change annualized without image-quality correction was examined after participants were combined by corresponding pattern label. Mean (SD) uncorrected change was −8.13 (12.46) mm³/y in principal C1, −34.00 (21.55) mm³/y in C2, and 16.79 (17.15) mm³/y in C3; median [IQR] values were −7.25 [−13.87 to −0.05], −30.32 [−43.10 to −21.27], and 13.87 [7.98 to 23.45] mm³/y, respectively. Thus, control-derived residualization did not create the low-change, pronounced-decrease, and increase directions; the same directional organization was evident in the uncorrected longitudinal volume data. This analysis was descriptive and was not used to redefine the principal center-specific assignments.

**Post-correction Diagnostic**

Among patients with complete longitudinal CAT12 image-quality data (n = 222), the post-correction association between longitudinal CAT12 image-quality change and mean image-quality-corrected annualized cortical-volume change was essentially null (Spearman rho = .0323; P = .6325). Mean (SD) longitudinal quality change was 0.0197 (0.2948), with median -0.0083; mean (SD) image-quality-corrected annualized cortical-volume change was -1.3993 (23.1360), with median -0.7852. Thus, after application of the control-derived correction, no detectable residual monotonic association remained between longitudinal image-quality change and mean image-quality-corrected annualized cortical-volume change.

**Pairwise Clinical Comparisons**

Exploratory pairwise Wilcoxon comparisons showed that, at baseline, functioning was higher in C1-P than C3-P (P = .0132), whereas negative-symptom severity was lower in C1-P than C3-P (P = .0209); the remaining baseline pairwise comparisons were not significant. At follow-up, functioning was lower in C2-P than C1-P (P = .0009) and C3-P (P = .0206), whereas C1-P and C3-P did not differ (P = .2276). Follow-up negative-symptom severity was greater in C2-P than C1-P (P = .0046); comparisons of C1-P with C3-P (P = .0808) and C2-P with C3-P (P = .1345) were not significant. Available sample sizes were C1-P/C2-P/C3-P = 77/66/79 for baseline functioning, 76/64/78 for baseline negative symptoms, 76/66/79 for follow-up functioning, and 77/66/79 for follow-up negative symptoms.

**Exploratory Clinical Comparison**

In the exploratory comparison of combined patient clusters 1 and 2 with patient cluster 3, nominal baseline differences in functioning and negative symptoms did not remain statistically significant after Holm adjustment. Follow-up outcomes did not differ, and nominally greater improvement in cluster 3 also did not remain significant after adjustment (eTable 5).

**Treatment and Compliance**

At follow-up, mean (SD) CPZ-equivalent dose was 192.7 (184.5) mg/d in patient cluster 1 (n = 72), 259.1 (186.2) mg/d in cluster 2 (n = 61), and 211.5 (198.7) mg/d in cluster 3 (n = 71; P = .06). Baseline compliance ratings were 5.66 (0.89) in cluster 1 (n = 76), 5.59 (0.84) in cluster 2 (n = 66), and 5.47 (0.99) in cluster 3 (n = 79; P = .51). Follow-up ratings were 5.28 (1.95), 5.62 (1.44), and 5.46 (1.70), respectively (n = 74, 65, and 76; P = .96). Among 213 patients with antipsychotic-regimen information at both visits, 150 (70.4%) changed named antipsychotic agent between V1 and V2. By cluster, 55 of 75 (73.3%) in cluster 1, 41 of 64 (64.1%) in cluster 2, and 54 of 74 (73.0%) in cluster 3 changed agent (P = .41) (eTable 6).

Treatment summaries used variable-specific available-case data; missing treatment observations were not imputed.

**Detailed Voxelwise Findings**

For the ordered-decrease contrast, the height threshold was T = 4.626517. The largest suprathreshold cluster included 118 739 voxels and peaked at T = 10.65 (Montreal Neurological Institute coordinates 4, 38, -2). Additional reported peaks were T = 6.20 at -12, 27, 52; T = 6.18 at 16, -52, -2; and T = 6.00 at 12, -60, 34 (eTable 7). The reverse contrast showed 1 cluster meeting the 50-voxel extent threshold (106 voxels; maximum T = 6.38), with 3 additional FWE-significant foci smaller than the extent threshold (28, 10, and 1 voxels).

The 4 peak coordinates (for clusters of 106, 28, 10, and 1 voxels, respectively) were 0, 9, 14; 2, -20, -3; 2, -32, 9; and 6, -27, 15. The Neuromorphometrics atlas labeled the 28-voxel focus as Right Ventral DC (46%), 3rd Ventricle (43%), Right Thalamus Proper (7%), and CSF (4%); atlas-overlap data were unavailable for the other 3 foci (eTable 8).

For patient cluster 3, the height threshold was T = 4.882235. The strongest clusters were in right ventral temporal cortex (7007 voxels; peak T = 8.71 at 30, -30, -24) and left ventral temporal cortex (4637 voxels; peak T = 8.07 at -48, -52, -14). The bilateral pattern included fusiform, inferior temporal, and parahippocampal cortex and extended to additional temporal, frontal, cingulate, subcortical, and cerebellar regions (eTable 12). The corresponding within-group decrease maps for control cluster 1 and patient clusters 1 and 2 are reported in eTables 9-11.

**Direct Ventricular-Volume Analysis**

Ventricular-volume/ICV change differed markedly across patient groups (ε² = .339; Holm-adjusted P < .001): mean changes were 0.0002 in C1-P, 0.0010 in C2-P, and −0.0006 in C3-P. V1 and V2 ratios did not differ after Holm adjustment (both P = .098; eTable 15).

**eTable 1. Baseline Characteristics Across Combined Center-Specific Patient Pattern Groups**

| **Characteristic** | **C1-P** | **C2-P** | **C3-P** | **P Value** | **Holm P Value** | **Test/Effect** |
| --- | --- | --- | --- | --- | --- | --- |
| Age, y | 29.33 (8.30); 27.65 [22.27, 33.56]; n = 77 | 28.58 (7.48); 27.10 [22.71, 33.07]; n = 66 | 26.52 (6.98); 25.11 [21.39, 29.86]; n = 79 | .0655 | .6845 | ε² .0244 |
| Sex, No. | Male 43; female 34 | Male 42; female 24 | Male 42; female 37 | .4274 | 1.0000 | χ² |
| BMI | 24.21 (4.65); 23.43 [21.05, 26.57]; n = 77 | 23.10 (3.02); 22.52 [20.92, 25.45]; n = 66 | 23.37 (3.60); 22.95 [20.63, 26.04]; n = 79 | .5754 | 1.0000 | ε² .0050 |
| Cigarettes/d | 12.90 (7.77); 12.50 [6.50, 20.00]; n = 21 | 10.57 (8.02); 10.00 [3.00, 20.00]; n = 27 | 8.84 (7.27); 10.00 [1.50, 15.00]; n = 24 | .2503 | 1.0000 | ε² .0124 |
| Education, No. (P/V/S/U) | 11/17/28/21 | 11/15/21/19 | 18/9/36/16 | .7614 | 1.0000 | KW |
| Diagnosis, No. (F20*/F23*/F25*) | 28/47/2 | 34/29/3 | 42/37/0 | .0498 | .5975 | Fisher exact |
| Age at onset, y | 28.94 (8.35); 27.40 [22.02, 33.30]; n = 77 | 28.15 (7.54); 26.69 [22.28, 32.84]; n = 66 | 26.07 (6.95); 24.99 [20.94, 29.58]; n = 79 | .0622 | .6845 | ε² .0249 |
| Illness duration at V1, y | 0.394 (0.391); 0.210 [0.155, 0.566]; n = 77 | 0.430 (0.392); 0.236 [0.128, 0.730]; n = 66 | 0.449 (0.422); 0.247 [0.136, 0.613]; n = 79 | .7946 | 1.0000 | ε² .0021 |
| DUP, mo | 1.42 (1.73); 1.00 [0.30, 1.50]; n = 77 | 2.52 (3.17); 1.00 [0.50, 3.00]; n = 66 | 1.69 (2.62); 1.00 [0.25, 1.75]; n = 79 | .1073 | .9658 | ε² .0200 |
| Baseline CPZ-equivalent dose, mg/d | 359.45 (224.60); 342.00 [188.00, 470.50]; n = 76 | 357.58 (185.12); 353.00 [255.00, 415.00]; n = 65 | 416.22 (252.69); 375.00 [245.25, 563.00]; n = 78 | .3185 | 1.0000 | ε² .0103 |
| Compliance score at baseline | 5.66 (0.89); 6 [5, 6]; n = 76 | 5.59 (0.84); 5.5 [5, 6]; n = 66 | 5.47 (0.99); 5 [5, 6]; n = 79 | .5051 | 1.0000 | KW |
| Baseline global GM/ICV ratio | 0.4406 (0.0263); 0.4369 [0.4218, 0.4528]; n = 77 | 0.4371 (0.0170); 0.4360 [0.4267, 0.4493]; n = 66 | 0.4384 (0.0261); 0.4363 [0.4210, 0.4569]; n = 79 | .9388 | 1.0000 | ε² .0006 |

***Note.*** *Values are mean (SD); median [IQR]; and available No. unless otherwise indicated. The diagnosis comparison used baseline diagnoses grouped as F20*/F23*/F25*.*

*Holm P values refer to adjustment across the prespecified 12 baseline comparisons. Holm-adjusted P values were .5975 for diagnosis, .6845 for age and age at onset, .9658 for DUP, and 1.0000 for the remaining 8 comparisons.*

***Abbreviations:*** *BMI, body mass index; CPZ, chlorpromazine equivalent; DUP, duration of untreated psychosis; GM/ICV, ratio of global gray matter volume to intracranial volume; ICD-10, International Statistical Classification of Diseases and Related Health Problems, Tenth Revision; IQR, interquartile range; KW, Kruskal-Wallis; P/V/S/U, primary, vocational, secondary, and university education; SD, standard deviation; V1, visit 1; ε², epsilon squared; χ², chi-square.*

**eTable 2. Center-Stratified Clinical Analyses**

| **Center** | **Time** | **Outcome** | **Cluster 1 / 2 / 3 means** | **Epsilon squared** | **Nominal P** |
| --- | --- | --- | --- | --- | --- |
| Site 1 | Baseline | Functioning | 71.97 / 67.00 / 71.12 | .0178 | .50 |
| Site 1 | Baseline | Negative symptoms | 2.28 / 2.45 / 2.33 | .0045 | .84 |
| Site 1 | Follow-up | Functioning | 83.48 / 68.21 / 82.73 | .1890 | <.001 |
| Site 1 | Follow-up | Negative symptoms | 1.89 / 2.64 / 2.02 | .1160 | .01 |
| Site 1 | Change | Functioning | 10.87 / 1.21 / 11.61 | .0606 | .09 |
| Site 1 | Change | Negative symptoms | -0.41 / 0.14 / -0.30 | .0530 | .12 |
| Site 2 | Baseline | Functioning | 72.76 / 66.50 / 60.91 | .0837 | .003 |
| Site 2 | Baseline | Negative symptoms | 1.96 / 2.20 / 2.69 | .0879 | .002 |
| Site 2 | Follow-up | Functioning | 79.38 / 72.96 / 75.24 | .0280 | .14 |
| Site 2 | Follow-up | Negative symptoms | 1.72 / 2.14 / 2.08 | .0389 | .06 |
| Site 2 | Change | Functioning | 6.62 / 6.46 / 14.33 | .0382 | .07 |
| Site 2 | Change | Negative symptoms | -0.24 / -0.06 / -0.61 | .0495 | .03 |

***Note.*** *Values are patient cluster-specific means from analyses performed separately in the 2 nonoverlapping imaging-center samples: site 1, MRI SITE 1 (n = 142 overall), and site 2, MRI SITE 2 (n = 258 overall). MRI SITE 1 center-specific clusters 1/2/3 contained 81/14/47 participants (patients, 32/14/33; controls, 49/0/14); MRI SITE 2 clusters contained 121/68/69 (patients, 45/52/46; controls, 76/16/23). Cluster 1, 2, and 3 labels denote within-center solutions; their average profiles were compared across centers in eTable 3, but participant-level assignment stability was not tested. Change is follow-up minus baseline; positive functioning change and negative negative-symptom change indicate improvement. Nominal P values are from 2-sided Kruskal-Wallis tests. Functioning was measured with the GAF scale, and negative symptoms with the consensus 5-factor PANSS negative-symptom score. GAF indicates Global Assessment of Functioning; PANSS, Positive and Negative Syndrome Scale; epsilon squared, effect size.*

**eTable 3. Cross-Center Similarity of Center-Specific Cluster Centroids**

| **Panel A. Observed Spearman Correlations Across 333 Parcels** | | | |
| --- | --- | --- | --- |
| **MRI SITE 1 cluster** | **MRI SITE 2 C1** | **MRI SITE 2 C2** | **MRI SITE 2 C3** |
| C1 | .1377 | .3110 | -.2189 |
| C2 | .3596 | .8646 | -.8320 |
| C3 | -.2166 | -.8441 | .8606 |
| **Panel B. Observed Unscaled Euclidean Distances Across 333 Parcels** | | | |
| **MRI SITE 1 cluster** | **MRI SITE 2 C1** | **MRI SITE 2 C2** | **MRI SITE 2 C3** |
| C1 | 102.9909 | 562.2387 | 650.7788 |
| C2 | 978.9781 | 480.8695 | 1599.4590 |
| C3 | 454.1499 | 1022.7313 | 251.3098 |
| **Panel C. Direct Matched-Centroid Correlations and Bootstrap Intervals** | | | |
| **Matched pair** | **Observed Spearman ρ** | **Bootstrap central estimate** | **95% interval** |
| MRI SITE 1 C1-MRI SITE 2 C1 | .1377 | .0881 | -.1486 to .3056 |
| MRI SITE 1 C2-MRI SITE 2 C2 | .8646 | .8005 | .7221 to .8520 |
| MRI SITE 1 C3-MRI SITE 2 C3 | .8606 | .7810 | .6867 to .8451 |
| **Panel D. C1-Referenced Direction-Vector Correlations** | | | |
| **Direction vector** | **Observed Spearman ρ** | **Bootstrap central estimate** | **95% interval** |
| C2 - C1 | .8228 | 0.7391 | 0.6244 to 0.8127 |
| C3 - C1 | .8122 | 0.7366 | 0.6288 to 0.8074 |

***Note****. A centroid is the mean 333-parcel image-quality-corrected annualized cortical-volume change profile of all participants assigned to a native center-specific cluster, irrespective of diagnosis. Panel A reports rank correlations across parcels; panel B reports unscaled Euclidean distances in the native centroid feature space. Panel C separates the observed correlations from the central estimates and 95% intervals in the 5000-resample bootstrap analysis. Panel D compares the C2 - C1 and C3 - C1 difference vectors across centers. Center-specific clusters were matched across centers according to centroid-profile similarity; panel B shows that each corresponding cluster pair was the closest Euclidean-distance pair, and panel A reports all rank correlations as complementary similarity summaries. C indicates cluster; MRI SITE 1, site 1; MRI SITE 2, site 2.*

**eTable 4. Imaging Site Composition of Combined Center-Specific Pattern Groups After Controls-Only Image-Quality Correction**

| **Imaging site** | **Combined C1** | **Combined C2** | **Combined C3** | **Total** |
| --- | --- | --- | --- | --- |
| MRI SITE 1 | 81 (57.0%) | 14 (9.9%) | 47 (33.1%) | 142 |
| MRI SITE 2 | 121 (46.9%) | 68 (26.4%) | 69 (26.7%) | 258 |
| **Total** | **202** | **82** | **116** | **400** |

***Note****. Values are No. (row %), except totals. Within each site and parcel, the unannualized V2-minus-V1 cortical-volume difference was modeled against longitudinal CAT12 image-quality change in controls. The full fitted value, including the intercept, was subtracted from each participant’s difference, and the residual was then divided by the interscan interval in years before clustering. Center-specific clusters were matched by centroid-profile similarity, and participants retained their center-specific assignments before being combined by corresponding pattern label. Site composition differed across the combined pattern groups (χ²₂ = 15.30, P < .001; Cramér V = 0.20), driven primarily by relative enrichment of MRI SITE 2 participants in C2. C1 and C3 had similar representation of the 2 imaging sites. C indicates cluster; MRI SITE 1, site 1; MRI SITE 2, site 2.*

**eTable 5. Exploratory Clinical Comparison of Combined Patient Clusters 1 and 2 With Patient Cluster 3**

| **Time point** | **Outcome** | **C1+C2** | **C3** | **P Value** | **Holm P Value** | **Direction** |
| --- | --- | --- | --- | --- | --- | --- |
| Baseline | GAF | 69.74 (15.70); 70 [60, 80]; n = 143 | 65.18 (15.99); 70 [55, 76.5]; n = 79 | .0303 | .1073 | C3 lower |
| Baseline | PANSS-N | 2.16 (0.81); 2.17 [1.50, 2.71]; n = 140 | 2.54 (1.03); 2.67 [1.67, 3.29]; n = 78 | .0102 | .0508 | C3 higher |
| Follow-up | GAF | 76.82 (14.03); 80 [69, 90]; n = 142 | 78.37 (12.82); 80 [75, 90]; n = 79 | .4961 | 1.0000 | No adjusted difference |
| Follow-up | PANSS-N | 2.00 (0.88); 1.83 [1.17, 2.67]; n = 143 | 2.05 (0.88); 1.83 [1.33, 2.67]; n = 79 | .6626 | 1.0000 | No adjusted difference |
| Change | GAF | 6.96 (16.11); 6.5 [-5, 18.75]; n = 142 | 13.19 (17.87); 10 [1, 23.5]; n = 79 | .0204 | .1022 | C3 greater increase |
| Change | PANSS-N | -0.17 (0.88); -0.17 [-0.67, 0.34]; n = 140 | -0.49 (1.12); -0.50 [-1.13, 0.17]; n = 78 | .0389 | .1556 | C3 greater decrease |

***Note.*** *Values are mean (SD); median [IQR]; and available No. C1+C2 combines patient groups defined by corresponding center-specific cluster labels 1 and 2; C3 denotes the patient group defined by corresponding center-specific cluster label 3. P values are from 2-sided Mann-Whitney U tests; Holm-adjusted P values are displayed. Change is follow-up minus baseline; positive GAF change and negative PANSS-N change indicate improvement.*

***Abbreviations:*** *GAF, Global Assessment of Functioning; IQR, interquartile range; PANSS-N, consensus 5-factor Positive and Negative Syndrome Scale negative-symptom score; SD, standard deviation.*

**eTable 6. Longitudinal Treatment and Compliance Indicators Across Combined Center-Specific Patient Pattern Groups**

| **Indicator** | **Cluster 1** | **Cluster 2** | **Cluster 3** | **P value** |
| --- | --- | --- | --- | --- |
| Baseline CPZ-equivalent dose, mean (SD), mg/d | 359.45 (224.60); n = 76 | 357.58 (185.12); n = 65 | 416.22 (252.69); n = 78 | .32 |
| Follow-up CPZ-equivalent dose, mean (SD), mg/d | 192.7 (184.5); n = 72 | 259.1 (186.2); n = 61 | 211.5 (198.7); n = 71 | .06 |
| Compliance score at baseline | 5.66 (0.89); n = 76 | 5.59 (0.84); n = 66 | 5.47 (0.99); n = 79 | .51 |
| Compliance score at follow-up | 5.28 (1.95); n = 74 | 5.62 (1.44); n = 65 | 5.46 (1.70); n = 76 | .96 |
| Changed antipsychotic agent between visits, No. (%) | 55/75 (73.3%) | 41/64 (64.1%) | 54/74 (73.0%) | .41 |

***Note.*** *Values are mean (SD) unless otherwise indicated. Cluster labels refer to patient groups formed by combining corresponding center-specific pattern labels. Visit-specific summaries used available-case data with variable-specific denominators; longitudinal medication-agent change was evaluated only among patients with nonmissing agent information at both visits. Missing treatment observations were not imputed. Medication compliance was rated on the 1-to-7 Hayward/Kemp scale, with higher values indicating better adherence.*

*Baseline CPZ-equivalent dose was available in 219 of 222 patients (1 missing observation in each combined center-specific patient pattern group); follow-up CPZ-equivalent dose was available in 204 patients; compliance was available in 221 patients at V1 and 215 at V2; and antipsychotic-agent change was assessed in 213 patients with agent information at both visits.*

**eFigure 1. Participant Flow Through the Study**

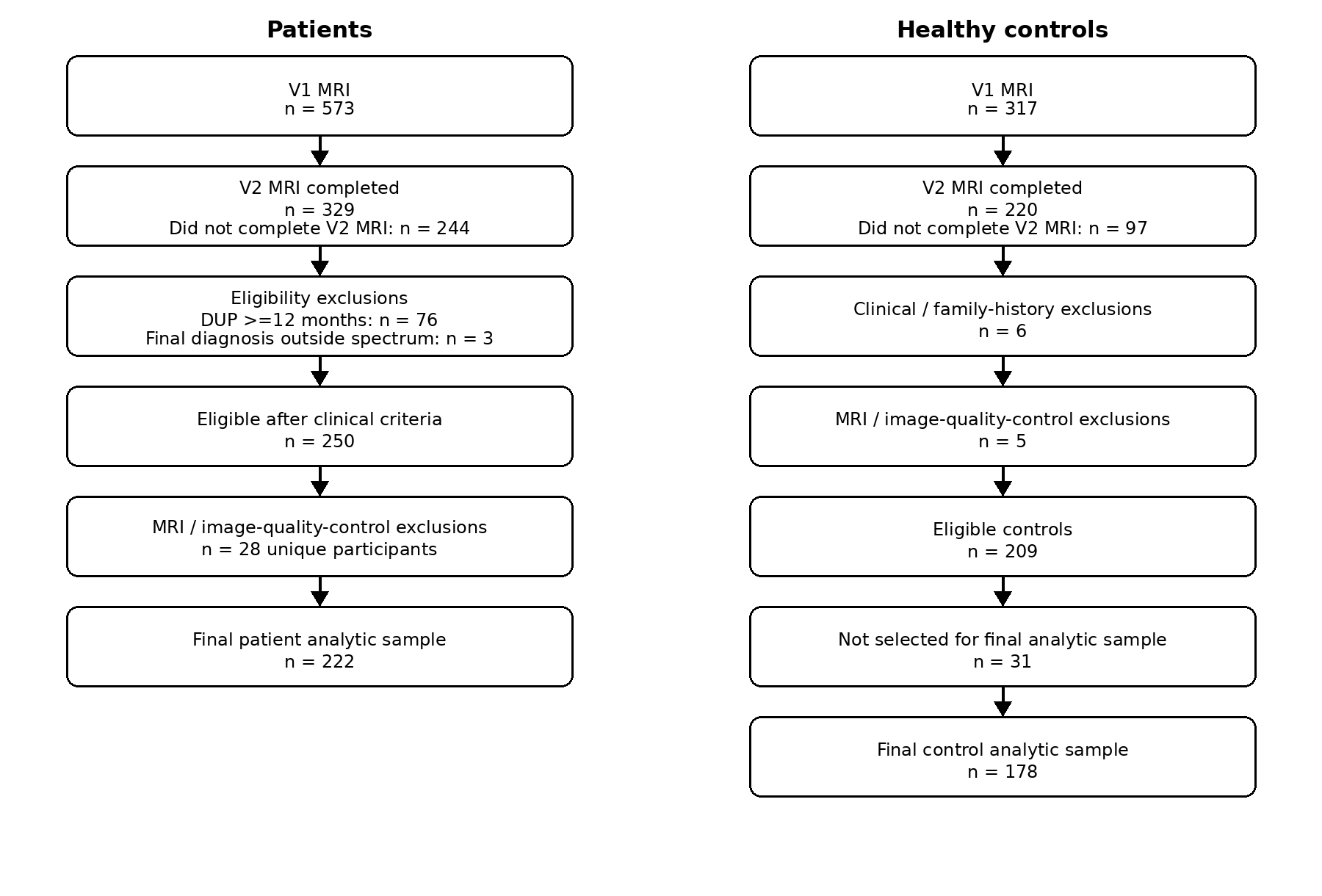

***Legend.*** *At dataset closure in April 2026, V1 MRI had been completed by 573 patients and 317 controls; V2 MRI was completed by 329 patients and 220 controls. In patients, 76 were excluded for DUP>=12 months and 3 for final diagnosis outside the prespecified spectrum, followed by 28 unique MRI/image-quality exclusions, yielding 222 patients. In controls, specified clinical/eligibility and MRI exclusions left 209 eligible controls, of whom 178 were frequency-matched to patients by site and baseline age; selection within overrepresented strata was independent of longitudinal cortical-volume change, image-quality change, and subsequent cluster assignment.*

**eFigure 2. Site-Specific Cluster Dendrograms**
A, Site 1 (MRI SITE 1). Diagnosis-blind Ward.D2 clustering of controls-only image-quality-corrected annualized cortical-volume changes (n = 142). The Cattell-Nelson-Gorsuch criterion selected K = 3 at both centers. The complementary Gap-statistic results are described in the eMethods; the Gap statistic also peaked at K = 3 at both centers, although differences among candidate solutions were modest.

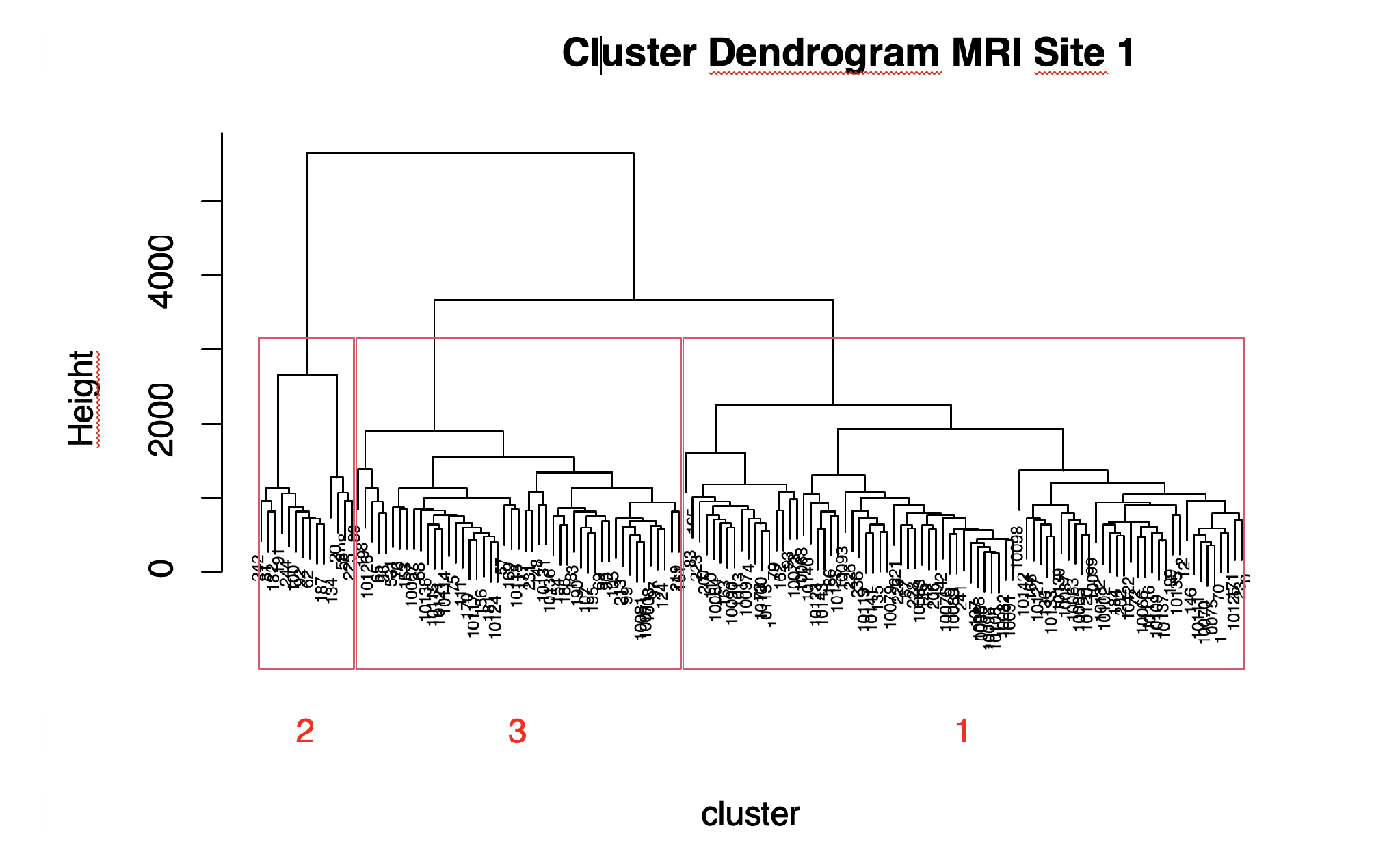

**B, Site 2 (MRI SITE 2).** Diagnosis-blind Ward.D2 clustering of controls-only image-quality-corrected annualized cortical-volume changes (n = 258). Red boxes indicate the Cattell-Nelson-Gorsuch 3-cluster solution.

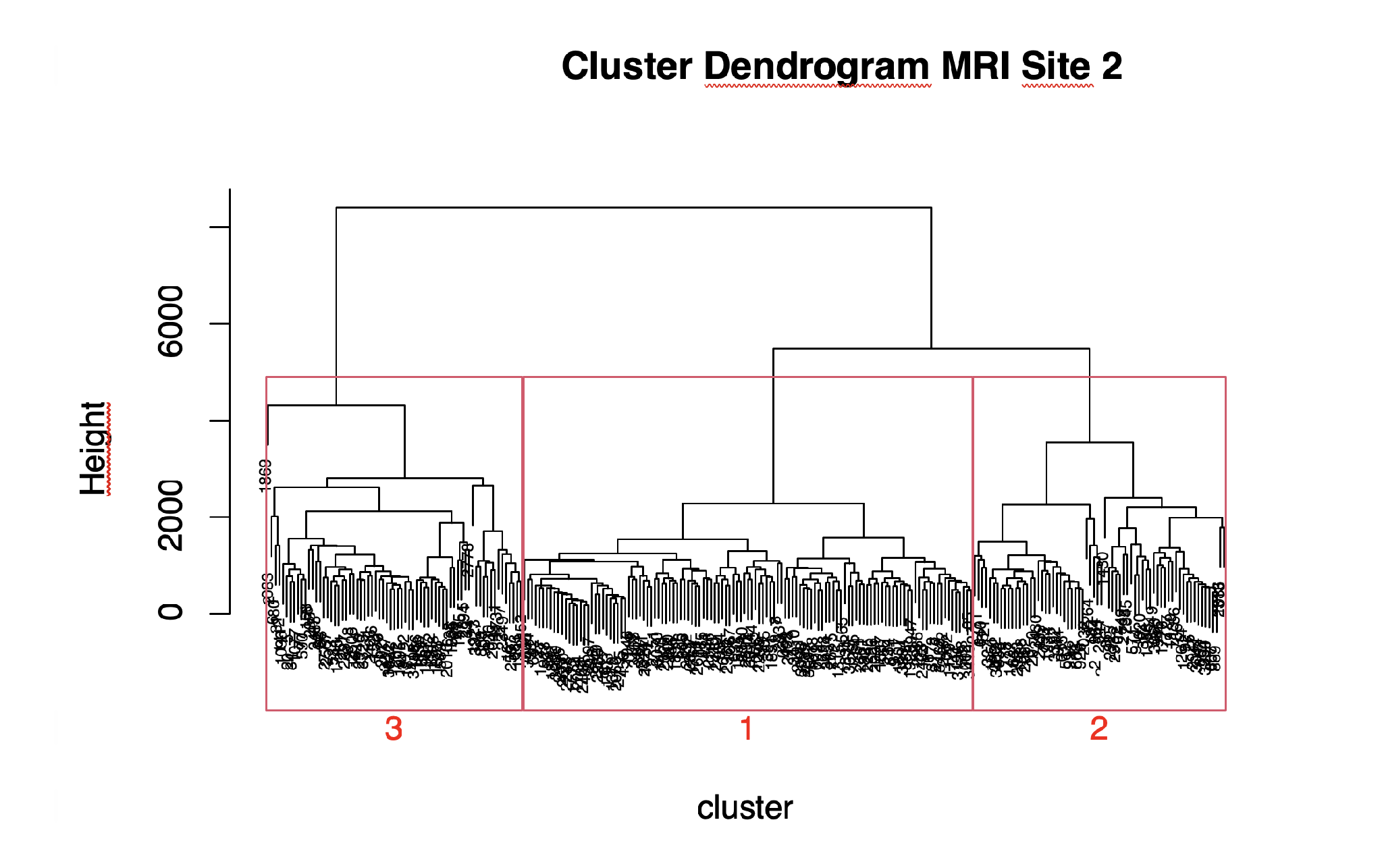

**eTable 7. Neuromorphometrics Atlas Labeling of Significant Clusters for the C1-HC–C1-P–C2-P Group Gradient Gray Matter Volume Decrease Contrast**

| **Cluster** | **Voxels** | **Peak T** | **Peak MNI x, y, z, mm** | **Neuromorphometrics Atlas Overlap** |
| --- | --- | --- | --- | --- |
| 1 | 118,739 | 10.65 | 4, 38, -2 | Right Cerebral White Matter (11%); Left Cerebral White Matter (9%); Right MTG middle temporal gyrus (5%); Right MFG middle frontal gyrus (3%); Left MTG middle temporal gyrus (3%); Right MSFG superior frontal gyrus medial segment (3%); Right AnG angular gyrus (3%); Left PCu precuneus (2%); Left AnG angular gyrus (2%); Right SFG superior frontal gyrus (2%); Right SMG supramarginal gyrus (2%); Right STG superior temporal gyrus (2%); Right ITG inferior temporal gyrus (2%); Left ACgG anterior cingulate gyrus (2%); Left MSFG superior frontal gyrus medial segment (2%); Right ACgG anterior cingulate gyrus (2%); Right SMC supplementary motor cortex (2%); Left MFG middle frontal gyrus (2%); Left SMC supplementary motor cortex (2%); Right MCgG middle cingulate gyrus (1%); Right PrG precentral gyrus (1%); Right AIns anterior insula (1%); Left SMG supramarginal gyrus (1%); Left ITG inferior temporal gyrus (1%); Left AIns anterior insula (1%); Right MOrG medial orbital gyrus (1%); Right TrIFG triangular part of the inferior frontal gyrus (1%); Right CO central operculum (1%); Left STG superior temporal gyrus (1%); Left MCgG middle cingulate gyrus (1%); Right FRP frontal pole (1%); Left SFG superior frontal gyrus (1%); Right OpIFG opercular part of the inferior frontal gyrus (1%) |
| 2 | 272 | 6.2 | -12, 27, 52 | Left SFG superior frontal gyrus (85%); Left Cerebral White Matter (15%) |
| 3 | 82 | 6.2 | 16, -52, -2 | Right LiG lingual gyrus (73%); Right Cerebral White Matter (24%); Right PCgG posterior cingulate gyrus (2%) |
| 4 | 75 | 6.1 | -51, 2, 24 | Left PrG precentral gyrus (59%); Left Cerebral White Matter (40%); Left OpIFG opercular part of the inferior frontal gyrus (1%) |
| 5 | 63 | 6 | 12, -60, 34 | Right Cerebral White Matter (76%); Right PCu precuneus (24%) |
| 6 | 624 | 5.9 | -15, -70, -26 | Left Cerebellum Exterior (91%); Left Cerebellum White Matter (6%); Left OFuG occipital fusiform gyrus (3%) |
| 7 | 59 | 5.8 | -21, -9, -20 | Left Amygdala (59%); Left Hippocampus (41%) |
| 8 | 531 | 5.8 | -15, -42, 64 | Left PoG postcentral gyrus (42%); Left Cerebral White Matter (27%); Left MPoG postcentral gyrus medial segment (16%); Left PCu precuneus (8%); Left SPL superior parietal lobule (7%) |
| 9 | 215 | 5.7 | 44, -62, -30 | Right Cerebellum Exterior (95%); Right ITG inferior temporal gyrus (5%) |
| 10 | 108 | 5.6 | -9, 12, 0 | Left Caudate (75%); Left Accumbens Area (13%); Left Cerebral White Matter (12%) |
| 11 | 113 | 5.6 | -36, -3, 46 | Left PrG precentral gyrus (50%); Left Cerebral White Matter (49%); Left MFG middle frontal gyrus (2%) |
| 12 | 96 | 5.6 | 6, -64, 8 | Right LiG lingual gyrus (40%); Right Calc calcarine cortex (39%); Right Cerebral White Matter (21%); Right Cun cuneus (1%) |
| 13 | 303 | 5.3 | -36, -76, -33 | Left Cerebellum Exterior (91%); Left Cerebellum White Matter (9%) |

***Note.*** *The table reports significant clusters from the negative contrast of the ordinal post hoc model coded control cluster 1, patient cluster 1, and patient cluster 2 as 0, 1, and 2, respectively (N = 268), testing increasingly negative annualized gray matter volume change. The model adjusted for age and sex. Significance was defined as voxel-level FWE-corrected P < .05 with a minimum extent of 50 contiguous voxels. Neuromorphometrics atlas regions comprising at least 1% of each cluster are displayed; percentages are rounded. Peak T is the maximum T statistic within each cluster. MNI indicates Montreal Neurological Institute; coordinates are in millimeters.*

**eFigure 3. Ordered C1-HC–C1-P–C2-P Gray Matter Volume Decrease**
Voxel-based morphometry map of increasingly negative annualized gray matter volume change across control cluster 1, patient cluster 1, and patient cluster 2, adjusted for age and sex; voxel-level FWE–corrected *P* < .05, minimum cluster extent 50 voxels
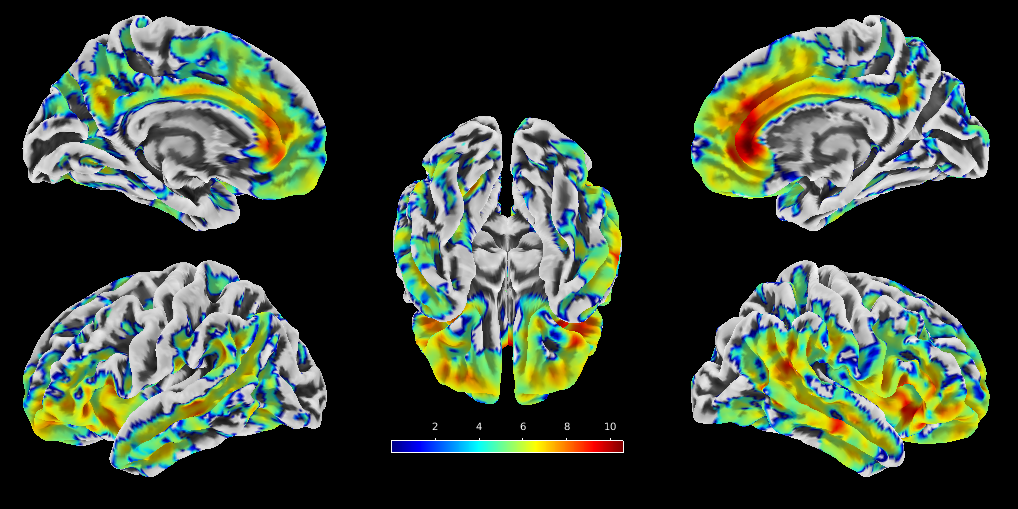

**eTable 8. Voxelwise Findings for the Reverse-Ordered C1-HC–C1-P–C2-P Group Gradient Gray Matter Volume Increase Contrast**

| **Cluster** | **Voxels** | **Peak T** | **Peak MNI x, y, z, mm** | **Neuromorphometrics Atlas Overlap** |
| --- | --- | --- | --- | --- |
| 1 | 106 | 6.38 | 0, 9, 14 | Atlas overlap unavailable |
| 2 | 28 | 6.23 | 2, -20, -3 | Right Ventral DC (46%); 3rd Ventricle (43%); Right Thalamus Proper (7%); CSF (4%) |
| 3 | 10 | 5.78 | 2, -32, 9 | Atlas overlap unavailable |
| 4 | 1 | 4.79 | 6, -27, 15 | Atlas overlap unavailable |

***Note.*** *The table reports positive effects for the reverse-ordered C1-HC–C1-P–C2-P group-gradient gray matter volume increase contrast. The ordinal model included control cluster 1, patient cluster 1, and patient cluster 2 (N = 268; 264 residual degrees of freedom) and adjusted for age and sex. The SPM analysis used a voxel-level FWE threshold of P < .05 (height threshold T = 4.626517) and displayed all clusters with display extent threshold k = 0. The prespecified reporting extent threshold remained 50 contiguous voxels; consequently, only the 106-voxel cluster met that combined criterion, while the 28-, 10-, and 1-voxel clusters were FWE significant but below the extent threshold. For clusters 1 through 4, respectively, cluster-level FWE-corrected P values were .004, .018, .030, and .045; cluster-level FDR-corrected q values were .300, .689, .780, and .891; cluster-level uncorrected P values were .075, .344, .585, and .891; peak-level FWE-corrected P values were .000, .000, .000, and .026; peak-level FDR-corrected q values were .002, .002, .011, and .515; peak Z values were 6.14, 6.01, 5.61, and 4.69; and peak-level uncorrected P values were .000 for all 4 clusters. Neuromorphometrics atlas overlap was available only for the 28-voxel cluster. MNI indicates Montreal Neurological Institute; coordinates are in millimeters.*

*Additional SPM output parameters: set-level P = .000 with c = 4; expected voxels per cluster, 33.817; expected number of clusters, .05; FWEp, 4.627; FDRp, 5.756; FWEc, 1; FDRc, Inf; FWHM, 15.1 x 14.8 x 14.7 mm (10.1 x 9.8 x 9.8 voxels); search volume, 1 291 248 mm3 (382 592 voxels; 295.1 resels); voxel size, 1.5 x 1.5 x 1.5 mm; and resel size, 972.93 voxels. The SPM table setting was 3 local maxima more than 8.0 mm apart.*

**eFigure 4. Reverse-Ordered C1-HC–C1-P–C2-P Gray Matter Volume Change**
Voxel-based morphometry map of the reverse ordered C1-HC–C1-P–C2-P contrast, adjusted for age and sex; voxel-level FWE–corrected *P* < .05. Only the 106-voxel cluster met the prespecified minimum extent of 50 voxels.
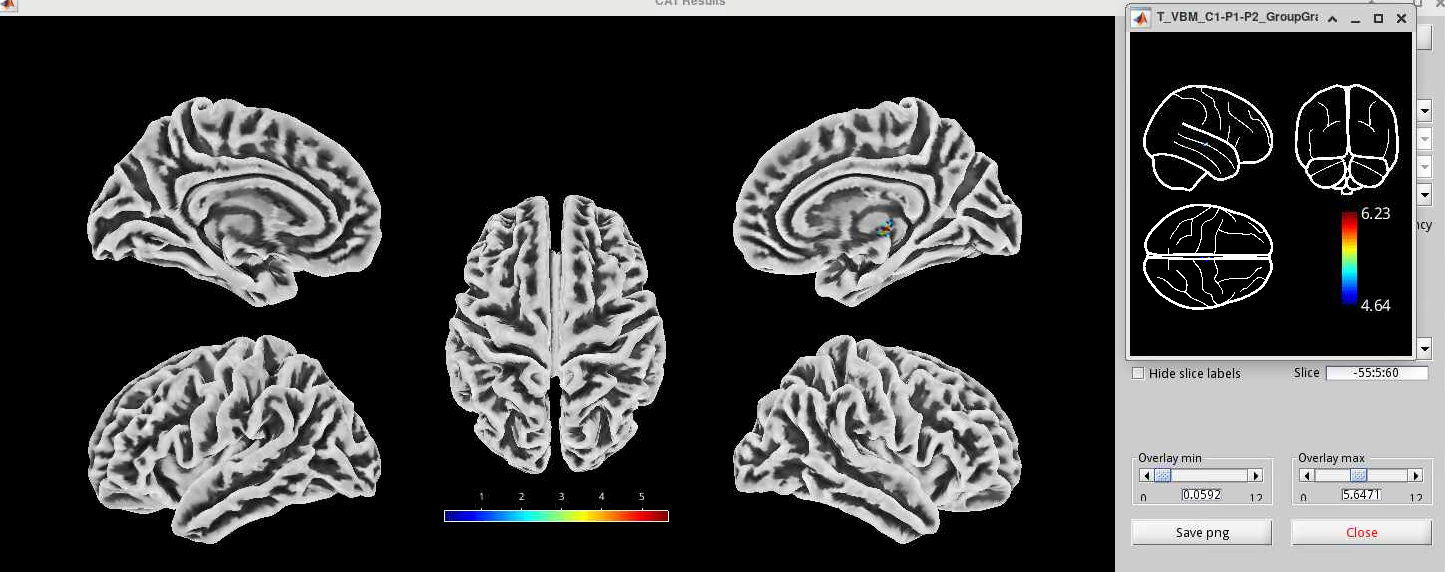

**eTable 9. Neuromorphometrics Atlas Labeling of Significant Clusters for Within-Group Decrease in Annualized Gray Matter Volume in Control Cluster 1**

| **Cluster** | **Voxels** | **Peak T** | **Peak MNI x, y, z, mm** | **Neuromorphometrics Atlas Overlap** |
| --- | --- | --- | --- | --- |
| 1 | 4,985 | 9.2 | 51, -48, 24 | Right AnG angular gyrus (37%); Right Cerebral White Matter (24%); Right MTG middle temporal gyrus (13%); Right SMG supramarginal gyrus (12%); Right STG superior temporal gyrus (9%); Right MOG middle occipital gyrus (3%); Right SPL superior parietal lobule (2%) |
| 2 | 8,560 | 8.9 | -45, -56, 18 | Left MTG middle temporal gyrus (24%); Left Cerebral White Matter (20%); Left AnG angular gyrus (19%); Left SMG supramarginal gyrus (15%); Left STG superior temporal gyrus (12%); Left MOG middle occipital gyrus (3%); Left SPL superior parietal lobule (2%); Left PO parietal operculum (2%); Left PT planum temporale (1%) |
| 3 | 5,483 | 8.1 | 9, -45, 42 | Right PCu precuneus (26%); Left PCu precuneus (23%); Right PCgG posterior cingulate gyrus (18%); Left PCgG posterior cingulate gyrus (17%); Right Cerebral White Matter (5%); Left MCgG middle cingulate gyrus (5%); Left Cerebral White Matter (4%); Right MCgG middle cingulate gyrus (3%) |
| 4 | 737 | 6.9 | 57, 9, -32 | Right TMP temporal pole (43%); Right MTG middle temporal gyrus (39%); Right ITG inferior temporal gyrus (11%); Right Cerebral White Matter (7%) |
| 5 | 337 | 6.9 | 12, 44, 9 | Right ACgG anterior cingulate gyrus (39%); Right MSFG superior frontal gyrus medial segment (33%); Right Cerebral White Matter (28%) |
| 6 | 1,225 | 6.8 | 32, 38, -14 | Right AOrG anterior orbital gyrus (30%); Right LOrG lateral orbital gyrus (23%); Right MFG middle frontal gyrus (15%); Right POrG posterior orbital gyrus (13%); Right Cerebral White Matter (10%); Right MOrG medial orbital gyrus (6%); Right FRP frontal pole (2%); Right SFG superior frontal gyrus (2%) |
| 7 | 414 | 6.7 | -8, 46, 3 | Left ACgG anterior cingulate gyrus (51%); Left MSFG superior frontal gyrus medial segment (43%); Left MFC medial frontal cortex (3%); Left Cerebral White Matter (2%) |
| 8 | 363 | 6.3 | -16, 15, 4 | Left Caudate (72%); Left Cerebral White Matter (28%) |
| 9 | 171 | 6.3 | 54, -36, 28 | Right Cerebral White Matter (41%); Right PO parietal operculum (33%); Right SMG supramarginal gyrus (23%); Right PT planum temporale (3%) |
| 10 | 271 | 6.2 | 18, -57, 24 | Right PCu precuneus (80%); Right Cerebral White Matter (20%) |
| 11 | 197 | 5.8 | -33, 26, -12 | Left OrIFG orbital part of the inferior frontal gyrus (38%); Left Cerebral White Matter (24%); Left POrG posterior orbital gyrus (14%); Left FO frontal operculum (12%); Left TrIFG triangular part of the inferior frontal gyrus (6%); Left AIns anterior insula (4%); Left LOrG lateral orbital gyrus (3%) |
| 12 | 240 | 5.8 | 36, 16, 6 | Right FO frontal operculum (69%); Right AIns anterior insula (21%); Right Cerebral White Matter (10%) |
| 13 | 91 | 5.7 | -27, 34, -15 | Left POrG posterior orbital gyrus (49%); Left MOrG medial orbital gyrus (30%); Left AOrG anterior orbital gyrus (15%); Left Cerebral White Matter (5%) |
| 14 | 116 | 5.6 | 45, -51, -12 | Right ITG inferior temporal gyrus (54%); Right Cerebral White Matter (30%); Right FuG fusiform gyrus (16%) |
| 15 | 153 | 5.6 | -30, 42, 22 | Left MFG middle frontal gyrus (88%); Left Cerebral White Matter (7%); Left SFG superior frontal gyrus (5%) |
| 16 | 305 | 5.5 | -33, 62, -14 | Left AOrG anterior orbital gyrus (47%); Left LOrG lateral orbital gyrus (20%); Left MFG middle frontal gyrus (20%); Left Cerebral White Matter (7%); Left SFG superior frontal gyrus (4%); Left FRP frontal pole (3%); Unknown (1%) |
| 17 | 144 | 5.3 | 46, 45, -2 | Right TrIFG triangular part of the inferior frontal gyrus (88%); Right MFG middle frontal gyrus (10%); Right Cerebral White Matter (3%) |

***Note.*** *The table reports significant clusters for within-group decrease in annualized gray matter volume in control cluster 1 (n = 125). The model adjusted for age and sex. Significance was defined as voxel-level FWE-corrected P < .05 with a minimum extent of 50 contiguous voxels. Neuromorphometrics atlas regions comprising at least 1% of each cluster are displayed; percentages are rounded. Peak T is the maximum T statistic within each cluster. MNI indicates Montreal Neurological Institute; coordinates are in millimeters.*

**eFigure 5. Gray Matter Volume Decrease in Control Cluster 1**
Within-group voxel-based morphometry map for control cluster 1 (n = 125), adjusted for age and sex; voxel-level–FWE-corrected *P* < .05, minimum cluster extent 50 voxels.

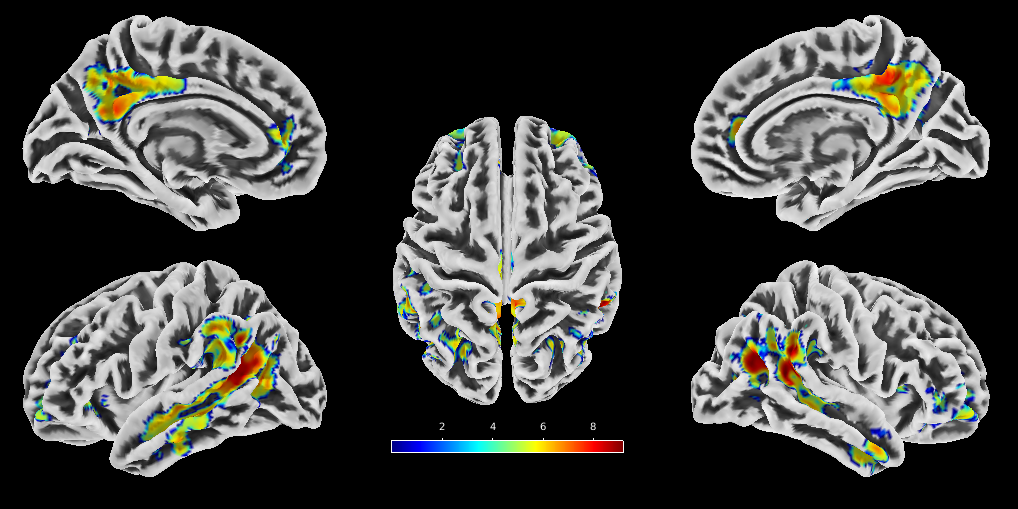

**eTable 10. Neuromorphometrics Atlas Labeling of Significant Clusters for Within-Group Decrease in Annualized Gray Matter Volume in Patient Cluster 1**

| **Cluster** | **Voxels** | **Peak T** | **Peak MNI x, y, z, mm** | **Neuromorphometrics Atlas Overlap** |
| --- | --- | --- | --- | --- |
| 1 | 18,449 | 9.5 | 22, 60, -6 | Right Cerebral White Matter (11%); Right ACgG anterior cingulate gyrus (9%); Left ACgG anterior cingulate gyrus (8%); Right MFG middle frontal gyrus (7%); Left Cerebral White Matter (6%); Right TrIFG triangular part of the inferior frontal gyrus (5%); Right MSFG superior frontal gyrus medial segment (5%); Right LOrG lateral orbital gyrus (4%); Right POrG posterior orbital gyrus (4%); Right AOrG anterior orbital gyrus (3%); Right FRP frontal pole (3%); Right OrIFG orbital part of the inferior frontal gyrus (3%); Left OrIFG orbital part of the inferior frontal gyrus (3%); Left AOrG anterior orbital gyrus (3%); Right MOrG medial orbital gyrus (3%); Left AIns anterior insula (2%); Left MFG middle frontal gyrus (2%); Right SFG superior frontal gyrus (2%); Left POrG posterior orbital gyrus (2%); Right FO frontal operculum (2%); Right OpIFG opercular part of the inferior frontal gyrus (2%); Right GRe gyrus rectus (2%); Left TrIFG triangular part of the inferior frontal gyrus (1%); Left FO frontal operculum (1%); Right MFC medial frontal cortex (1%); Left MSFG superior frontal gyrus medial segment (1%); Left LOrG lateral orbital gyrus (1%); Left FRP frontal pole (1%); Right AIns anterior insula (1%) |
| 2 | 8,978 | 9.5 | 50, -48, 28 | Right AnG angular gyrus (27%); Right MTG middle temporal gyrus (22%); Right Cerebral White Matter (21%); Right STG superior temporal gyrus (10%); Right SMG supramarginal gyrus (9%); Right PO parietal operculum (3%); Right PoG postcentral gyrus (2%); Right SPL superior parietal lobule (1%); Right CO central operculum (1%) |
| 3 | 6,127 | 9 | -50, -51, 33 | Left MTG middle temporal gyrus (27%); Left AnG angular gyrus (25%); Left Cerebral White Matter (19%); Left STG superior temporal gyrus (12%); Left SMG supramarginal gyrus (11%); Left ITG inferior temporal gyrus (3%); Left PT planum temporale (1%); Left PO parietal operculum (1%) |
| 4 | 2,285 | 8.1 | -8, -48, 33 | Left PCu precuneus (29%); Right PCu precuneus (25%); Left PCgG posterior cingulate gyrus (17%); Right PCgG posterior cingulate gyrus (16%); Right Cerebral White Matter (9%); Left Cerebral White Matter (5%) |
| 5 | 1,113 | 6.9 | 10, -21, 40 | Left MCgG middle cingulate gyrus (48%); Right MCgG middle cingulate gyrus (26%); Right MPrG precentral gyrus medial segment (9%); Right Cerebral White Matter (7%); Left SMC supplementary motor cortex (5%); Left Cerebral White Matter (2%); Right SMC supplementary motor cortex (1%) |
| 6 | 465 | 6.7 | -12, 9, 12 | Left Caudate (80%); Left Cerebral White Matter (11%); Left Lateral Ventricle (9%) |
| 7 | 111 | 6.5 | -46, 4, -28 | Left Cerebral White Matter (48%); Left TMP temporal pole (42%); Left MTG middle temporal gyrus (5%); Left STG superior temporal gyrus (5%) |
| 8 | 597 | 6.5 | 54, -2, -36 | Right MTG middle temporal gyrus (60%); Right ITG inferior temporal gyrus (19%); Right Cerebral White Matter (11%); Right TMP temporal pole (6%); Right STG superior temporal gyrus (4%) |
| 9 | 297 | 6.3 | 44, 15, 39 | Right MFG middle frontal gyrus (88%); Right Cerebral White Matter (10%); Right PrG precentral gyrus (3%) |
| 10 | 60 | 6.1 | 27, -72, 27 | Right Cerebral White Matter (50%); Right SOG superior occipital gyrus (37%); Right SPL superior parietal lobule (13%) |
| 11 | 241 | 6.1 | -9, -42, 50 | Left PCu precuneus (73%); Left MPrG precentral gyrus medial segment (12%); Left PCgG posterior cingulate gyrus (6%); Left Cerebral White Matter (4%); Left MPoG postcentral gyrus medial segment (4%) |
| 12 | 147 | 5.9 | -51, -9, -30 | Left Cerebral White Matter (39%); Left MTG middle temporal gyrus (33%); Left ITG inferior temporal gyrus (28%) |
| 13 | 105 | 5.9 | 10, 15, 33 | Right MCgG middle cingulate gyrus (42%); Right Cerebral White Matter (30%); Right SMC supplementary motor cortex (28%) |
| 14 | 93 | 5.9 | 12, -33, 51 | Right MPrG precentral gyrus medial segment (74%); Right Cerebral White Matter (15%); Right MPoG postcentral gyrus medial segment (4%); Right PCu precuneus (4%); Right PCgG posterior cingulate gyrus (2%) |
| 15 | 144 | 5.8 | 34, -21, 16 | Right PIns posterior insula (41%); Right Cerebral White Matter (30%); Right PO parietal operculum (15%); Right TTG transverse temporal gyrus (14%) |
| 16 | 58 | 5.5 | -50, -27, 33 | Left SMG supramarginal gyrus (48%); Left Cerebral White Matter (33%); Left PoG postcentral gyrus (19%) |

***Note.*** *The table reports significant clusters for within-group decrease in annualized gray matter volume in patient cluster 1 (n = 77). The model adjusted for age and sex. Significance was defined as voxel-levelFWE-corrected P < .05 with a minimum extent of 50 contiguous voxels. Neuromorphometrics atlas regions comprising at least 1% of each cluster are displayed; percentages are rounded. Peak T is the maximum T statistic within each cluster. MNI indicates Montreal Neurological Institute; coordinates are in millimeters.*

**eFigure 6. Gray Matter Volume Decrease in Patient Cluster 1**
Within-group voxel-based morphometry map for patient cluster 1 (n = 77), adjusted for age and sex; voxel-level

FWE–corrected *P* < .05, minimum cluster extent 50 voxels.

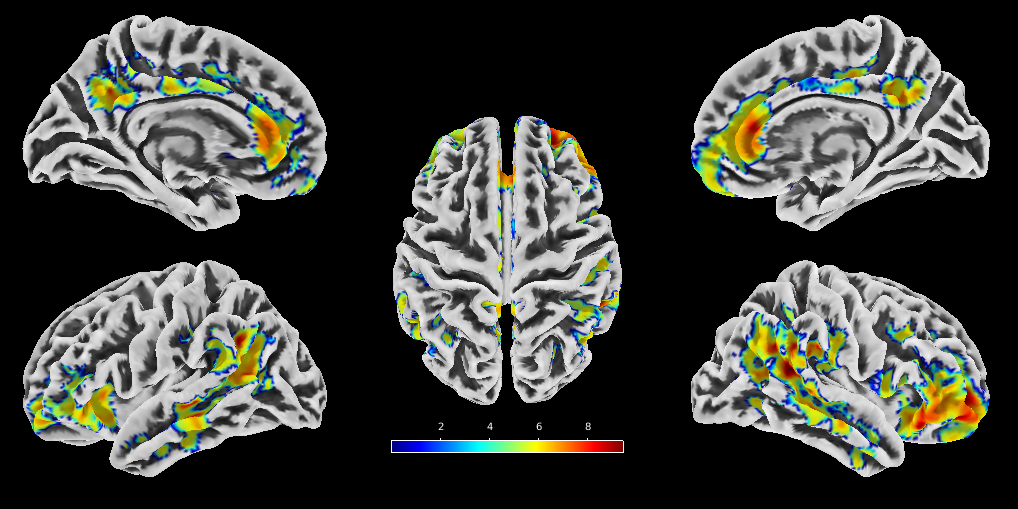

**eTable 11. Neuromorphometrics Atlas Labeling of Significant Clusters for Within-Group Decrease in Annualized Gray Matter Volume in Patient Cluster 2**

| **Cluster** | **Voxels** | **Peak T** | **Peak MNI x, y, z, mm** | **Neuromorphometrics Atlas Overlap** |
| --- | --- | --- | --- | --- |
| 1 | 157,921 | 12 | 48, -44, 24 | Right Cerebral White Matter (10%); Left Cerebral White Matter (9%); Right MTG middle temporal gyrus (4%); Right MFG middle frontal gyrus (4%); Left MTG middle temporal gyrus (3%); Left MFG middle frontal gyrus (3%); Left PCu precuneus (2%); Left AnG angular gyrus (2%); Right AnG angular gyrus (2%); Right MSFG superior frontal gyrus medial segment (2%); Right PCu precuneus (2%); Left SMG supramarginal gyrus (2%); Right SFG superior frontal gyrus (2%); Right ITG inferior temporal gyrus (2%); Left SFG superior frontal gyrus (2%); Left ITG inferior temporal gyrus (2%); Right SMG supramarginal gyrus (2%); Right STG superior temporal gyrus (2%); Left ACgG anterior cingulate gyrus (1%); Right ACgG anterior cingulate gyrus (1%); Right MCgG middle cingulate gyrus (1%); Left STG superior temporal gyrus (1%); Left MSFG superior frontal gyrus medial segment (1%); Right SMC supplementary motor cortex (1%); Left MCgG middle cingulate gyrus (1%); Left MOrG medial orbital gyrus (1%); Left SMC supplementary motor cortex (1%); Right PrG precentral gyrus (1%); Right TMP temporal pole (1%); Right MOrG medial orbital gyrus (1%); Left AIns anterior insula (1%) |
| 2 | 1,791 | 7.4 | 15, -86, -34 | Right Cerebellum Exterior (95%); Right Cerebellum White Matter (5%) |
| 3 | 1,790 | 7 | -15, -72, -26 | Left Cerebellum Exterior (89%); Left Cerebellum White Matter (10%); Left OFuG occipital fusiform gyrus (1%) |
| 4 | 71 | 6.6 | -21, -8, -20 | Left Amygdala (65%); Left Hippocampus (35%) |
| 5 | 596 | 6.5 | -14, -26, 8 | Left Thalamus Proper (100%) |
| 6 | 68 | 6.4 | 4, -57, -50 | Right Cerebellum Exterior (100%) |
| 7 | 98 | 6.3 | 28, -87, 20 | Right SOG superior occipital gyrus (96%); Right Cerebral White Matter (2%); Right MOG middle occipital gyrus (2%) |
| 8 | 227 | 6.1 | 10, -12, 9 | Right Thalamus Proper (100%) |
| 9 | 464 | 6.1 | 15, -84, 39 | Right Cun cuneus (41%); Right SOG superior occipital gyrus (29%); Right Cerebral White Matter (29%) |
| 10 | 168 | 5.9 | 27, -50, 64 | Right SPL superior parietal lobule (88%); Right Cerebral White Matter (12%) |
| 11 | 76 | 5.8 | 15, -100, 10 | Right OCP occipital pole (74%); Right Cerebral White Matter (24%); Right Cun cuneus (3%) |

***Note.*** *The table reports significant clusters for within-group decrease in annualized gray matter volume in patient cluster 2 (n = 66). The model adjusted for age and sex. Significance was defined as voxel-level FWE-corrected P < .05 with a minimum extent of 50 contiguous voxels. Neuromorphometrics atlas regions comprising at least 1% of each cluster are displayed; percentages are rounded. Peak T is the maximum T statistic within each cluster. MNI indicates Montreal Neurological Institute; coordinates are in millimeters.*

**eFigure 7. Gray Matter Volume Decrease in Patient Cluster 2**
Within-group voxel-based morphometry map for patient cluster 2 (n = 66), adjusted for age and sex; voxel-level

FWE–corrected *P* < .05, minimum cluster extent 50 voxels.

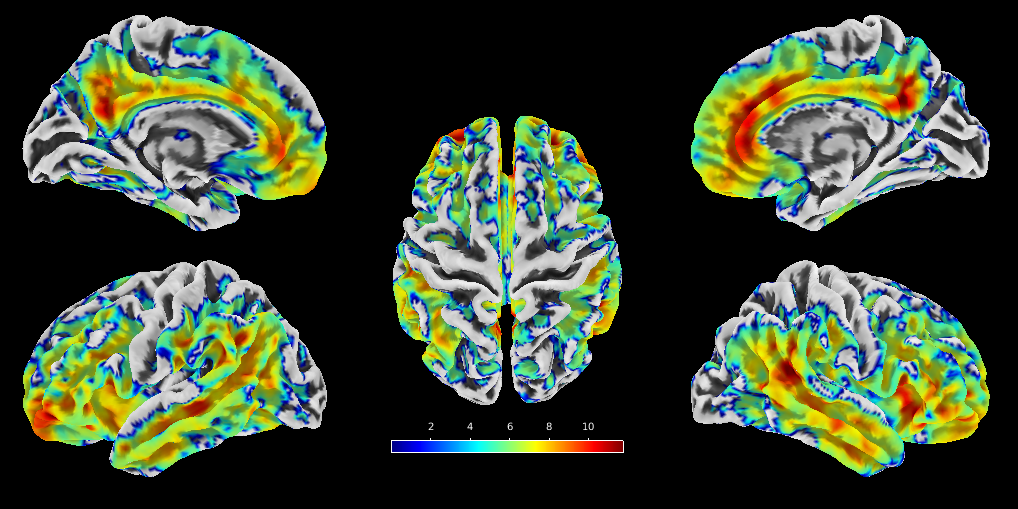

**eTable 12. Neuromorphometrics Atlas Labeling of Significant Clusters for Within-Group Increase in Annualized Gray Matter Volume in Patient Cluster 3**

| **Cluster** | **Voxels** | **Peak T** | **Peak MNI x, y, z, mm** | **Neuromorphometrics Atlas Overlap** |
| --- | --- | --- | --- | --- |
| 1 | 7,007 | 8.7 | 30, -30, -24 | Right FuG fusiform gyrus (28%); Right ITG inferior temporal gyrus (17%); Right Cerebral White Matter (16%); Right Cerebellum Exterior (9%); Right Thalamus Proper (6%); Right PHG parahippocampal gyrus (6%); Right LiG lingual gyrus (5%); Right OFuG occipital fusiform gyrus (5%); Right IOG inferior occipital gyrus (3%); Right Hippocampus (2%); Right MTG middle temporal gyrus (2%) |
| 2 | 4,637 | 8.1 | -48, -52, -14 | Left FuG fusiform gyrus (36%); Left Cerebral White Matter (21%); Left ITG inferior temporal gyrus (16%); Left Cerebellum Exterior (9%); Left PHG parahippocampal gyrus (6%); Left LiG lingual gyrus (3%); Left OFuG occipital fusiform gyrus (3%); Left MTG middle temporal gyrus (3%); Left Cerebellum White Matter (2%); Left Hippocampus (2%) |
| 3 | 1,795 | 8 | 33, -6, -6 | Right Cerebral White Matter (51%); Right Putamen (34%); Right Pallidum (13%); Right AIns anterior insula (1%) |
| 4 | 720 | 7.5 | -12, -52, -54 | Left Cerebellum Exterior (90%); Left Cerebellum White Matter (10%) |
| 5 | 1,139 | 7.2 | 54, -12, 2 | Right Cerebral White Matter (29%); Right PP planum polare (23%); Right TMP temporal pole (20%); Right STG superior temporal gyrus (11%); Right TTG transverse temporal gyrus (10%); Right CO central operculum (4%); Right PT planum temporale (3%) |
| 6 | 644 | 7 | -51, 2, -10 | Left TMP temporal pole (33%); Left Cerebral White Matter (28%); Left PP planum polare (22%); Left STG superior temporal gyrus (16%); Left TTG transverse temporal gyrus (2%) |
| 7 | 561 | 6.8 | 14, -52, -52 | Right Cerebellum Exterior (94%); Right Cerebellum White Matter (6%) |
| 8 | 317 | 6.7 | -16, -26, 0 | Left Thalamus Proper (90%); Left Cerebral White Matter (8%); Left Ventral DC (2%) |
| 9 | 526 | 6.7 | 52, 16, 20 | Right OpIFG opercular part of the inferior frontal gyrus (75%); Right Cerebral White Matter (20%); Right MFG middle frontal gyrus (5%) |
| 10 | 408 | 6.3 | -40, 2, 27 | Left PrG precentral gyrus (51%); Left Cerebral White Matter (30%); Left OpIFG opercular part of the inferior frontal gyrus (11%); Left MFG middle frontal gyrus (9%) |
| 11 | 334 | 6.3 | -3, -28, 34 | Left PCgG posterior cingulate gyrus (58%); Left Cerebral White Matter (31%); Left MCgG middle cingulate gyrus (10%) |
| 12 | 1,059 | 6.2 | -30, -6, -10 | Left Putamen (48%); Left Cerebral White Matter (40%); Left Pallidum (7%); Left Accumbens Area (4%) |
| 13 | 112 | 6.1 | -26, -2, -33 | Left Ent entorhinal area (79%); Left Cerebral White Matter (20%); Left TMP temporal pole (2%) |
| 14 | 86 | 5.6 | 26, -62, -27 | Right Cerebellum Exterior (97%); Right Cerebellum White Matter (3%) |
| 15 | 176 | 5.6 | 40, 8, 52 | Right MFG middle frontal gyrus (86%); Right Cerebral White Matter (14%) |
| 16 | 85 | 5.4 | 60, -20, 3 | Right Cerebral White Matter (53%); Right STG superior temporal gyrus (47%) |

***Note.*** *The table reports significant clusters for within-group increase in annualized gray matter volume in patient cluster 3 (n = 79). The model adjusted for age and sex. Significance was defined as voxel-level FWE-corrected P < .05 with a minimum extent of 50 contiguous voxels. Neuromorphometrics atlas regions comprising at least 1% of each cluster are displayed; percentages are rounded. Peak T is the maximum T statistic within each cluster. MNI indicates Montreal Neurological Institute; coordinates are in millimeters.*

**eFigure 8. Gray Matter Volume Increase in Patient Cluster 3**
Within-group voxel-based morphometry map for patient cluster 3 (n = 79), adjusted for age and sex; voxel-level FWE–corrected *P* < .05, minimum cluster extent 50 voxels.

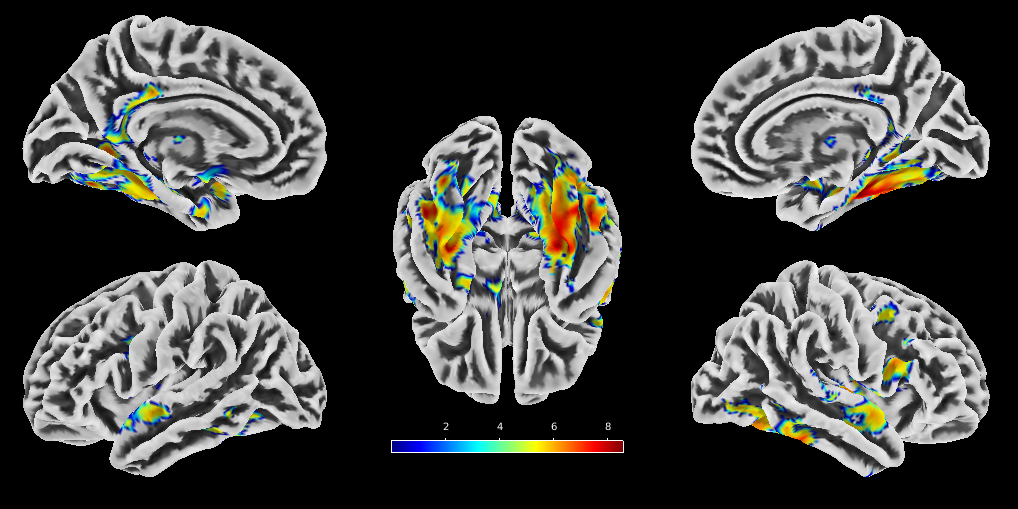

**eTable 13. Image-Quality Correction and Sensitivity Analyses**

| **Analysis** | **Center-specific cluster number** | **Combined corresponding pattern sizes** | **Patient-control allocation** | **Follow-up GAF** | **Follow-up PANSS-N** | **Residual quality association** |
| --- | --- | --- | --- | --- | --- | --- |
| Controls-only calibration | MRI SITE 1: 3; MRI SITE 2: 3 | 202 / 82 / 116 | P < .001 | P = .0010; Holm P = .005 | P = .0043; Holm P = .02 | rho = .0323; P = .6325 |
| All-participant sensitivity | MRI SITE 1: 4; MRI SITE 2: 3 | Not applicable; K differed by center | Not reported | Not reported | Not reported | Not evaluated |

Note. Principal analysis used the controls-only calibration. The residual quality association shown for the principal analysis was calculated among patients with complete longitudinal CAT12 image-quality data (n = 222; Spearman rho = .0323; P = .6325). Because K differed by center in the all-participant sensitivity analysis, cross-center matched pattern groups were not defined; pooled patient-control and clinical P values are therefore not reported.

**eTable 14. Uncorrected Cortical-Volume Change Sensitivity Checks**

| **Analysis / Measure** | **Low-change / C1** | **Pronounced-decrease / C2** | **Increase / C3** |
| --- | --- | --- | --- |
| **Principal combined corresponding pattern groups, No.** | 202 | 82 | 116 |
| Controls/patients, No. | 125/77 | 16/66 | 37/79 |
| Corresponding uncorrected change, mean (SD), mm³/y | −8.13 (12.46) | −34.00 (21.55) | 16.79 (17.15) |
| Corresponding uncorrected change, median [IQR], mm³/y | −7.25 [−13.87 to −0.05] | −30.32 [−43.10 to −21.27] | 13.87 [7.98 to 23.45] |

***Note.*** *Uncorrected values are annualized cortical-volume changes without CAT12 image-quality correction. Uncorrected values within the principal combined corresponding pattern groups showed the same low-change, pronounced-decrease, and increase directions. No inferential tests were performed because cluster membership was data-derived.*

**eTable 15. Ventricular Volume Normalized to Intracranial Volume Across Patient Pattern Groups**

| ***Measure*** | ***C1-P*** | ***C2-P*** | ***C3-P*** | ***P Value*** | ***Holm P Value*** | ***ε²*** |
| --- | --- | --- | --- | --- | --- | --- |
| *V1 ventricular volume/ICV* | *0.0099 (0.0033); 0.0094 [0.0078 to 0.0118]; n = 77* | *0.0111 (0.0040); 0.0102 [0.0084 to 0.0131]; n = 66* | *0.0114 (0.0041); 0.0105 [0.0084 to 0.0138]; n = 79* | *.0928* | *.0982* | *.0213* |
| *V2 ventricular volume/ICV* | *0.0101 (0.0033); 0.0098 [0.0077 to 0.0120]; n = 77* | *0.0121 (0.0046); 0.0107 [0.0087 to 0.0150]; n = 66* | *0.0109 (0.0040); 0.0098 [0.0081 to 0.0127]; n = 79* | *.0491* | *.0982* | *.0270* |
| *V2 minus V1 change* | *0.0002 (0.0006); 0.0001 [-0.0003 to 0.0005]; n = 77* | *0.0010 (0.0013); 0.0008 [0.0003 to 0.0015]; n = 66* | *-0.0006 (0.0009); -0.0004 [-0.0010 to 0.0000]; n = 79* | *<.001* | *<.001* | *.3390* |

***Note.*** *Values are mean (SD); median [IQR]; and available No. Ventricular volume was normalized to ICV. Change is V2 minus V1. P values are from Kruskal-Wallis tests; Holm adjustment was applied across the V1, V2, and change comparisons. All pairwise post hoc comparisons of longitudinal change were P < .001. ICV, intracranial volume; IQR, interquartile range; SD, standard deviation; V1, visit 1; V2, visit 2.*

**Additional Descriptive Statistics for V2 Minus V1 Change**

| **Patient pattern group** | **Minimum** | **Maximum** | **Missing values, No.** |
| --- | --- | --- | --- |
| C1-P | -0.0013 | 0.0028 | 0 |
| C2-P | -0.0013 | 0.0063 | 0 |
| C3-P | -0.0031 | 0.0011 | 0 |

*Note. Values are dimensionless changes in ventricular volume/ICV. The source reports no missing values in any patient pattern group.*

**eFigure 9. Ventricular-Volume/ICV Change Across Patient Pattern Groups**

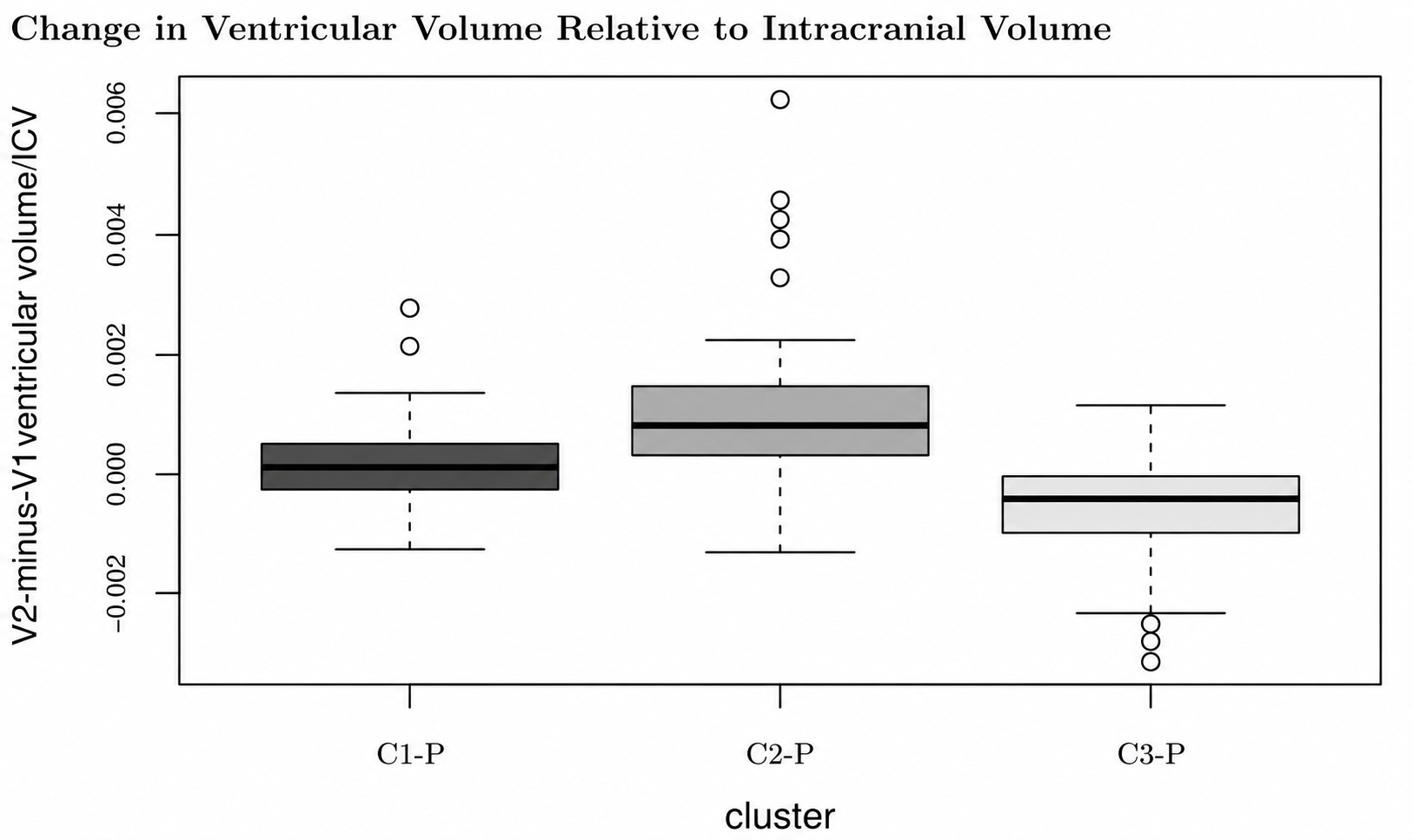

*Legend. Original boxplot of V2-minus-V1 change in ventricular volume normalized to intracranial volume (dKomoryICV; dimensionless). Source cluster labels 1, 2, and 3 correspond to C1-P, C2-P, and C3-P, respectively. Descriptive statistics and inferential comparisons are reported in eTable 15.*

**eTable 16. Annualized Cortical-Volume Change by Imaging Center, Diagnosis, and Pattern Group**

| **Group** | **Minimum** | **1st quartile** | **Median** | **Mean** | **3rd quartile** | **Maximum** | **SD** | **N** |
| --- | --- | --- | --- | --- | --- | --- | --- | --- |
| **Panel A. IKEM (MRI SITE 1)** | | | | | | | | |
| C1-HC | -17.6077 | -4.6707 | -1.3466 | -1.4785 | 2.1795 | 12.3208 | 6.0991 | 49 |
| C1-P | -18.8432 | -8.6650 | -3.1853 | -2.8376 | 2.5160 | 14.7711 | 7.6385 | 32 |
| C2-HC |  |  |  |  |  |  |  | 0 |
| C2-P | -73.8956 | -52.4372 | -30.7240 | -38.3265 | -25.2912 | -20.0197 | 19.0558 | 14 |
| C3-HC | 4.9843 | 6.8617 | 13.5539 | 14.4829 | 18.4635 | 41.6882 | 9.4739 | 14 |
| C3-P | 4.1145 | 9.0256 | 15.5665 | 15.9120 | 19.9265 | 40.6750 | 7.9522 | 33 |
| **Panel B. NUDZ (MRI SITE 2)** | | | | | | | | |
| C1-HC | -15.9686 | -6.1935 | -1.8222 | -1.3981 | 3.1714 | 12.1502 | 6.5665 | 76 |
| C1-P | -18.0453 | -7.8012 | -1.3790 | -2.1103 | 3.6555 | 11.5552 | 7.8044 | 45 |
| C2-HC | -34.0459 | -20.5941 | -15.4632 | -16.5071 | -11.7547 | -4.5315 | 8.5085 | 16 |
| C2-P | -66.7062 | -29.2330 | -22.0023 | -23.5378 | -15.0059 | -4.0198 | 13.1494 | 52 |
| C3-HC | 6.7049 | 9.9267 | 14.9102 | 17.4192 | 22.6765 | 40.3089 | 8.9877 | 23 |
| C3-P | 3.3175 | 15.7501 | 20.0897 | 24.1429 | 27.2774 | 111.2595 | 17.4352 | 46 |
| **Panel C. Combined centers** | | | | | | | | |
| C1-HC | -17.6077 | -5.2990 | -1.5109 | -1.4296 | 3.0813 | 12.3208 | 6.3625 | 125 |
| C1-P | -18.8432 | -8.4059 | -1.8152 | -2.4125 | 3.3100 | 14.7711 | 7.6936 | 77 |
| C2-HC | -34.0459 | -20.5941 | -15.4632 | -16.5071 | -11.7547 | -4.5315 | 8.5085 | 16 |
| C2-P | -73.8956 | -34.5564 | -23.2266 | -26.6748 | -16.8959 | -4.0198 | 15.6653 | 66 |
| C3-HC | 4.9843 | 9.5529 | 14.8966 | 16.3082 | 21.4272 | 41.6882 | 9.1575 | 37 |
| C3-P | 3.3175 | 13.4130 | 18.4096 | 20.7047 | 22.7295 | 111.2595 | 14.7651 | 79 |

*Note. Values summarize mean cortical-volume change after controls-only image-quality correction and subsequent annualization (mm³/y). C1, C2, and C3 denote corresponding center-specific pattern groups; HC, healthy controls; P, patients; SD, standard deviation; N, number of observations.*

**Data Sharing Statement**

**Data available:** Yes.

**Data types:** Deidentified participant-level data and data dictionary.

**Data shared:** Participant ID, patient/control status, imaging site, cluster assignment, and image-quality-corrected annualized cortical-volume change across 333 parcels.

**Supporting documents:** Analytic code.

**When available:** With publication.

**How to access data:** Zenodo (DOI: <https://doi.org/10.5281/zenodo.23211633>).

**Who can access the data:** Anyone.

**Types of analyses:** Any research purpose.

**Restrictions:** Raw MRI and additional potentially identifying clinical data will not be publicly shared.
